# Clinical Characteristics and Outcomes of Critically ill Mechanically Ventilated COVID-19 Patients Receiving interleukin-6 Receptor Antagonists and Corticosteroid Therapy: A Multicenter International Observational Study

**DOI:** 10.1101/2021.04.12.21255323

**Authors:** Marwa Amer, Mohammed Bawazeer, Khalid Maghrabi, Ahmed M. Kamel, Abid Butt, Talal Dahhan, Eiad Kseibi, Syed Moazzum Khurshid, Mohammed Abujazar, Razan Alghunaim, Muath Rabee, Maal Abualkhair, Ali Al-Janoubi, Abeer Turki AlFirm, Ognjen Gajic, Allan J. Walkey, Jarrod M Mosier, Igor Borisovich Zabolotskikh, Oscar Y Gavidia, Santiago Y. Teruel, Michael A. Bernstein, Karen Boman, Vishakha K. Kumar, Vikas Bansal, Rahul Kashyap

## Abstract

**Objectives:** The interest in interleukin-6 receptor antagonists (IL-6RA) and steroids have increased recently due to their potential role as immunomodulatory effect in critically ill coronavirus disease (COVID-19). The magnitude of this therapy in subgroups of patients with invasive mechanical ventilation (MV) remains to be fully clarified. We compared the clinical characteristics and outcomes of patients requiring iMV, and receiving IL-6RA and steroids with different steroids regimens.

**Design:** International, multicenter, observational study derived from Viral Infection and Respiratory Illness University Study registry and conducted through Discovery Network, Society of Critical Care Medicine. Marginal structural modeling was used to adjust time-dependent confounders; observations were weighted using inverse probability of treatment weight. A sensitivity analysis was conducted for target trial design.

**Setting:** 168 hospitals, 16 countries.

**Patients:** Covid-19 ICU patients (≥18 years) requiring MV between March 01,2020, and January 10,2021.

**Intervention:** None.

**Measurements and Main Results:** Of 860 patients met eligibility criteria, 589 received steroids, 170 IL-6RA, and 101 combinations; groups were balanced after adjustment. Median daily steroid dose was 7.5 mg dexamethasone or equivalent (IQR:6-14 mg); 80.8% and 19.2% received low-dose and high-dose steroids, respectively. The median C-reactive protein level was >75 mg/L in majority of our cohort. The use of IL-6R antagonists alone or in combination was not associated with a significant difference in ventilator-free days (VFD) compared to steroids alone with different steroids regimens (adjusted incidence rate ratio [95% CI]): IL-6R antagonists (1.12 [0.88,1.4]), combination (0.83 [0.6,1.14]). Patients treated with low or high-dose steroids had non-significant differences in VFD compared to IL-6RA (ß=0.62, 95% CI −1.54,2.78 for low-dose steroid; ß=-1.19, 95% CI −3.85,1.47 for high-dose steroid). There was no difference in 28-day mortality and hospital mortality with IL-6RA alone or in combination compared to steroids alone (28-day mortality adjusted odds ratio [95% CI]): IL-6RA (0.68[0.44,1.07]), combination (1.07[0.67,1.70]). Sensitivity analysis findings were consistent with primary analysis. Liver dysfunction was higher in IL-6RA (p=0.04) while rate of bacteremia did not differ among groups.

**Conclusions:** In adult ICU COVID-19 patients on iMV, we found no difference in outcomes between those who received IL-6RA, steroids, or combination therapy and those who received IL-6RA or low-or high-dose steroids. Further randomized trials are needed to enhance our understanding for IL-6RA safety with different steroids regimen and the magnitude of benefit in those subgroups of patients.

## Background

Severe coronavirus disease 2019 (COVID-19) can manifest as respiratory failure with elevated inflammatory markers, resulting in cytokine storms, for which interleukin-6 receptor antagonists (IL-6RA) are approved treatments (1–3). Interest in IL-6RA and corticosteroids has increased recently due to their potential role as immunomodulators (4–7). In view of the results from International Randomized, Embedded, Multi-factorial, Adaptive Platform Trial for Community-Acquired Pneumonia (REMAP-CAP) and Randomized Evaluation of COVID-19 Therapy (RECOVERY), guidelines by National Institutes of Health and Infectious Disease Society of America conditionally suggest tocilizumab in combination with steroids (low-dose dexamethasone, 6 mg daily for 10 days) for ICU patients exhibiting rapid respiratory failure progression or high inflammatory markers (8–11). Data with other steroid regimens are limited (high-dose dexamethasone, 20 mg IV daily) which are commonly used for non-COVID-19 acute respiratory distress syndrome (ARDS) and may be utilized for COVID-19 ARDS (12, 13). Moreover, a limited number of patients on invasive mechanical ventilation (MV) were included in the previous trials (29 % in REMAP-CAP, 14 % in RECOVERY, 16 % in TOCIBRAS, and 37% in COVACTA) (13). Therefore, the magnitude of this combination therapy in subgroups of patients with iMV remains to be fully clarified and was identified as a research priority to enhance personalized clinical decision-making (14, 15). We sought to describe clinical characteristics and outcomes of ICU patients requiring iMV, and receiving IL-6RA and steroids using the data from the largest COVID-19 registry and provided further insights into sarilumab and utilization pattern of multiple doses of steroids.

## Materials and Methods

### Data Source

Data for this ancillary study were derived from Viral Infection and Respiratory Illness Universal Study (VIRUS) registry, an international, multicenter, observational study conducted through Discovery Network, Society of Critical Care Medicine (SCCM). The study was approved by SCCM Scientific Review Committee, SCCM Discovery Steering Committee, and King Faisal Specialist Hospital and Research Center (KFSH&RC) Institutional Review Board (IRB) (IRB# 2201053, registered at ClinicalTrials.gov NCT04486521). In some participating hospitals, verbal consent was obtained whereas in other countries consent was waived by local research ethics committees. Local investigators were responsible for obtaining local approval in line with applicable regulations. VIRUS is one of largest registries that consecutively collects data on COVID-19 patients. Details of this database described elsewhere (16–19) (Supplemental Table 1). The study is reported following the STrengthening the Reporting of Observational Studies in Epidemiology (STROBE) and the Risk Of Bias In Nonrandomized Studies of Interventions (ROBINS-I) guidelines (16–19).

### Study Participants and Exposures

Adult patients (≥ 18 years) were eligible if admitted to ICU from March 01, 2020 to January 10, 2021, required invasive mechanical ventilation (iMV), had positive polymerase chain reaction (PCR) SARS-CoV-2, and received IL-6RA (tocilizumab or sarilumab), corticosteroid (dexamethasone, hydrocortisone, methylprednisolone, or prednisone), or combination. We excluded patients who were < 17 years, repeatedly admitted to ICU during the same hospital visit, on chronic systemic steroid at home or taking steroids for indications other than COVID-19, or died before receiving IL-6RA or steroid. We stratified population into three groups: IL-6RA, corticosteroid, and combination. Corticosteroid group was stratified further based on dexamethasone equivalent (mg) into high and low-dose. High-dose was defined as > 15 mg/day of dexamethasone or equivalent (12, 13). Patients were followed until hospital discharge, death, 28 days, or January 13th, 2021-date on which database was locked for analysis.

### Data Collection

The following variables were collected on days 0 (ICU admission), 1-3, 7, 14, 21, and 28 from electronic medical records according to standard operating procedure: demographics, biomarkers (ferritin, interleukin-6 (IL-6), D-dimer, fibrinogen, C-reactive protein [CRP], lactate dehydrogenase, and lymphocyte count), microbiology, concomitant medications, MV duration, discharge status, PaO2(mm Hg)/FiO2 (arterial partial pressure of oxygen over fractional inspired oxygen concentration; PF ratio), and timing of drug initiation and dose. Data were collected online and stored on a secure data server using Mayo Clinic Research Electronic Data Capture application (REDCap). Data quality was assessed routinely to ensure completeness and accuracy.

### Study Outcomes and Variable Definitions

Primary outcome was ventilator-free days score (VFD) from iMV at day 28 which was chosen as a patient-centered outcome and highly influenced by mortality (20). Secondary outcomes included: ICU and hospital mortality, 28-day mortality, hospital and ICU length of stay (LOS), median change in PF ratio, adverse events (AEs), and biomarkers concentration. Details on outcome definitions are provided in Supplemental Table 2.

### Statistical Analysis

Sample size was determined pragmatically, based on all available ICU patients in VIRUS database who met eligibility criteria. Statistical analyses were performed using R software, V3.6.3 (Vienna, Austria). Counts and percentages were used to represent categorical variables. Continuous data were summarized using means ± standard deviations (SDs) or medians (interquartile ranges). Chi-square test was used to compare distribution of categorical variables and Kruskal–Wallis test and ANOVA were used to compare distributions of continuous non-normal and normal variables, respectively.

We evaluate three statistical models for this study. Model one was for Marginal structural models (MSM) in the primary analysis which included the overall treatment strategy not solely restricted to the first 48 hours. Model two was exploratory analyses with added Sequential Organ Failure Assessment (SOFA) score, and highest FiO2 to the base model. Model three is a sensitivity analysis to emulate target trial approach restricted to the treatment received in the first 48 hours.

#### Model 1

Marginal structural models (MSM) were used to compare three regimens after adjusting for non-time and time-varying covariates (21). Observations were weighted by inverse probability of treatment weight (IPTW). Nonparametric modeling using generalized boosted model (GBM) estimate IPTWs with automatic handling of missing data (22). TWANG package in R was used for analysis. Covariates distributions were compared before and after applying propensity weights. Standardized mean difference (SMD) was used to assess balance of pseudo-population using Kolmogorov–Smirnov stop rule. Average treatment effect (ATE) estimation method was used throughout analysis on entire sample. Covariates included in model were judged as likely to influence outcomes and have been identified in several studies: age; sex; ethnicity; asthma/chronic obstructive pulmonary disease (COPD); ARDS grade; history of diabetes, hypertension, coronary heart disease, or congestive heart failure; the lowest FiO2; therapeutic anticoagulation; hydroxychloroquine; azithromycin; and antivirals (including remdesivir); vasopressors; and paralytics (**Supplemental File 1**). Analyses were performed using robust variance estimators taking into account clustering within hospitals and patients. Quasi-Poisson generalized linear modeling was used to compare VFD after weighting. Patients who died before 28-day were assigned zero VFD. Quasi-Poisson regression coefficients were exponentiated to obtain incidence rate ratio (IRR), which is expected change in VF-days (ratio), compared to reference category. Quasibinomial generalized linear modeling was used to compare mortality between three groups. Weighted Kaplan–Maier estimator was used to compare hospital and ICU LOS. Patients who were still in ICU after 28-day study period were censored at 28 days. Log-rank test was used to compare survival before and after weighing. Cause-Specific Cox regression analysis was used to assess factors associated with mortality at 28 days. Discharge and mortality were included as competing risks. Linear mixed modeling was used to compare change in laboratory values and PF ratio. Hypothesis testing was performed at 5% level of significance.

#### Model 2

In exploratory analyses of potential factors associated with variation in VFD and mortality, we added Sequential Organ Failure Assessment (SOFA) score, and highest FiO2 to base model 1.

#### Model 3

We performed a sensitivity analysis to emulate target trial to reduce immortal time bias. Eligible patients for target trial approach were those on MV, received IL-6RA or steroids within ICU days 0 or 1. Patients who died within first 48 hours were excluded. Those who get treatment of interest after day 1 were classified by treatment exposure in first 48 hours (similar to intention-to-treat analysis in randomized trial) (23). The outcomes of interest in target trial approach were VFD and 28-day mortality.

## Results

### Overall Characteristics

A total of 23,783 patients were screened, 860 met eligibility criteria and classified as follows: 170 received IL-6RA, 589 received steroids, and 101 received both therapies. In sensitivity analysis to emulate target trial, 562 patients fulfilled inclusion criteria: 406 received steroids (72.2%), 121 received IL6-RA (21.5%), and 35 received both (6.23%) (Figure 1). In unadjusted analysis, baseline characteristics were not balanced. IL-6RA patients were younger (median 57.5 years vs. 61.1 in steroid vs. 61.8 in combination; p=0.009) and chronic pulmonary diseases, excluding asthma (COPD, bronchiectasis, and interstitial lung disease), were more prevalent in steroid group (59 [10.0%] vs. 7 [4.12%] in IL-6RA vs. 16 [15.8%] in combination, p=0.005). The ARDS was prevalent in 35.5% of steroid-treated patients, 14.7% in the IL-6R antagonist, and 36.6 % in combination therapy; the majority of these patients had severe ARDS grades (PF ratio < 100). There was no significant difference in the incidence of acute liver failure, and bacteremia among the three arms. After adjustments for baseline covariates, SMD was ∼0.1 for majority of the covariates, indicating three groups were more likely balanced (Table 1).

**Figure 1.**
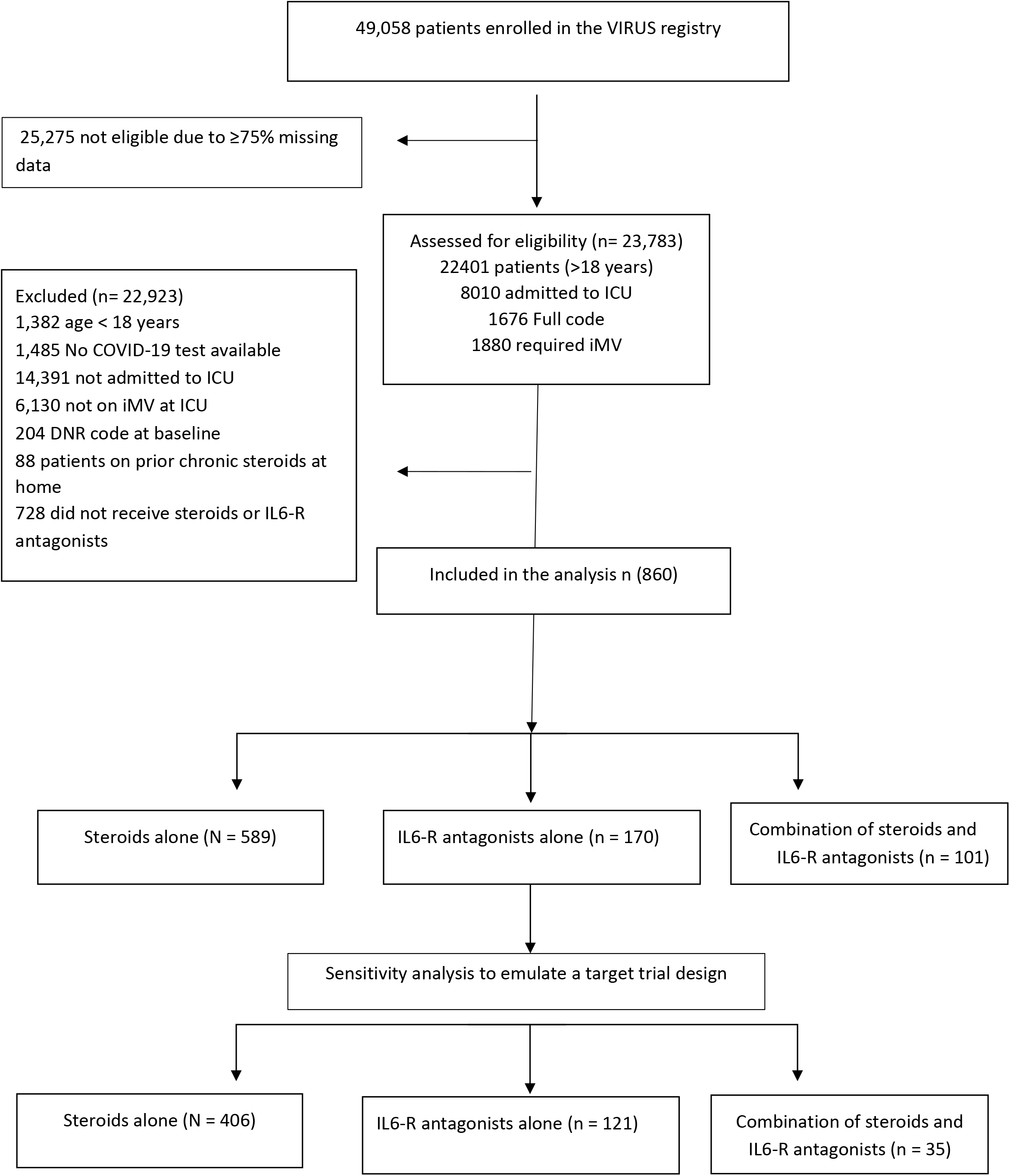
Flow chart for data extraction from VIRUS database. DNR = Do-not-resuscitate order; iMV = invasive mechanical ventilation; VIRUS = Viral Infection and Respiratory Illness University Study registry

**Table 1.** Baseline characteristics after adjustments for baseline covariates

| Baseline characteristics | Steroids | IL6-R antagonists | Both | P | SMD |
| --- | --- | --- | --- | --- | --- |
| N<sup>a</sup> | 811.6 | 596.8 | 698.9 |  |  |
| Male | 528 (65.1) | 430 (72.1) | 433 (62) | 0.25 | 0.145 |
| Age (mean (SD)) | 60.9 (14.2) | 60.6 (13.8) | 61.5 (12.8) | 0.90 | 0.042 |
| Weight at admission (mean (SD)) | 91.3 (27.9) | 92.7 (26.3) | 94.5 (29.9) | 0.62 | 0.076 |
| Ethnic group |  |  |  | 0.93 | 0.113 |
| Hispanic | 163 (20.1) | 128 (21.5) | 147 (21.3) |  |  |
| Non-Hispanic | 385 (47.4) | 285 (47.8) | 325 (47) |  |  |
| CVD | 176 (21.6) | 75 (12.6) | 161 (23) | 0.11 | 0.183 |
| Asthma/COPD | 143 (17.6) | 84 (14.1) | 147 (21) | 0.33 | 0.121 |
| Hydroxychloroquine | 164 (20.2) | 181 (30.2) | 180 (26) | 0.15 | 0.141 |
| Azithromycin | 300 (37) | 279 (46.7) | 294 (42.1) | 0.32 | 0.122 |
| Antiviral | 145 (18) | 78 (13.1) | 141 (20.2) | 0.42 | 0.136 |
| ARDS grade |  |  |  | 0.79 | 0.148 |
| No | 542 (66.8) | 441 (74) | 489 (70) |  |  |
| Mild (P:F 200-300) | 22 ( 2.8) | 17 (3) | 23 ( 3.3) |  |  |
| Moderate (P:F 100-199) | 83 (10.7) | 38 ( 6.5) | 41 ( 6) |  |  |
| Severe (P:F< 100) | 132 (16.9) | 79 (13.8) | 113 (16.2) |  |  |
| Any Anticoagulant | 693 (85.3) | 504 (84.4) | 560 (80.1) | 0.46 | 0.093 |
| Therapeutic anticoagulation | 179 (22.1) | 87 (14.6) | 162 (23.1) | 0.15 | 0.147 |
| FiO2 lowest (mean (SD)) | 0.56 (0.25) | 0.52 (0.24) | 0.57 (0.24) | 0.43 | 0.127 |
| Neuromuscular blocker | 659 (82) | 520 (87) | 625 (89.4) | 0.15 | 0.16 |
| Vasopressors | 638 (78.6) | 506 (84.8) | 612 (87.5) | 0.14 | 0.16 |

Data presented as n (%) unless otherwise specified. N (%) may differ due to approximation
<sup>a</sup> Each individual gets own weight that is used for further analysis. Multiplication by these weights can usually results in decimals.
ARDS = Acute respiratory distress syndrome; CVD = cardiovascular diseases; COPD = Chronic obstructive pulmonary disease; SMD = standardized mean difference; SD = standard deviation; P:F = ratio of arterial oxygen partial pressure (PaO2 in mmHg) to fractional inspired oxygen

The median daily steroid dose was 7.5 mg dexamethasone equivalent (IQR 6-14 mg). Of these, 80.8% received low-dose steroids, and 19.2% received high-dose steroids. Of the patients who received IL-6RA, the majority (84.5%) received only one dose of tocilizumab while second and third doses were administered to 13.1% and 2.3%, respectively. Similarly, most patients (84.7%) in the sarilumab group received a single dose. The dosage range of tocilizumab was 600–800 mg and sarilumab was 200-400 mg (**Table 2**). Matched patients received comparable amounts of sedatives, paralytics, and anticoagulation. The CRP level (assessed in 413) was consistently > 75 mg/L across all groups while IL-6 concentrations (assessed in 24) were highest in combination group. Additional comorbidities, ventilator and radiology characteristics, ICU supports, and biomarker levels are included in Supplemental Tables 3-11.

**Table 2.** Utilization pattern of steroids and IL6-R antagonists in relation to ICU admission

| **Dose and timing of administration** | **Value** |
| --- | --- |
| **Average daily dose (mg dexamethasone equivalent) <sup>a</sup>** | 7.50 [6, 14.1] |
| Low steroid dose | 496 (80.8) |
| High steroid dose <sup>b</sup> | 118 (19.2) |
| **Date of steroid start in relation to ICU admission <sup>c</sup>** |  |
| 0 | 384 (55.6) |
| 1 | 102 (14.8) |
| 2 | 37 (5.4) |
| 3 | 22 (3.2) |
| 4 | 24 (3.5) |
| After day 5 | 118 (17) |
| **Tocilizumab** | 213 (78.3) |
| **Tocilizumab number of doses <sup>d</sup>** |  |
| 1 | 180 (84.5) |
| 2 | 28 (13.1) |
| 3 | 5 (2.3) |
| **Sarilumab** | 59 (21.7) |
| **Sarilumab number of doses <sup>d</sup>** |  |
| 1 | 50 (84.7) |
| 2 | 6 (10) |
| >2 | 3 (5.1) |
| **Date of IL6-R antagonists start in relation to ICU** |  |
| 0 | 129 (39.5) |
| 1 | 55 (20.3) |
| 2 | 30 (11.1) |
| 3 | 21 (7.8) |
| 4 | 4 (1.5) |
| After day 5 | 32 (11.8) |

<sup>a</sup> The dose was available to 614 patients. The average daily dose of steroids was calculated by averaging the daily doses received by the patients during the ICU stay
<sup>b</sup> High dose was defined as > 15 mg/day of dexamethasone or dexamethasone equivalent
<sup>c</sup> The proportion was calculated from the number of patients who received the steroids (n = 693).
<sup>d</sup> The proportion was calculated from the number of patients who received IL6-R antagonists intravenous formulation (n=272).

### Primary Outcomes

The results for outcomes before adjusting for covariates is presented in Supplemental Tables 12. After adjustment (model 1), we found no significant difference in VFDs between the use of IL-6R antagonists alone or in combination compared to using steroids alone with different steroids regimen (adjusted IRR [95% CI]): IL-6RA (1.12 [0.88,1.40]), combination (0.83 [0.6,1.14]). Exploratory analyses findings (model 2) were comparable to primary analysis. Sensitivity analysis as target trial design (model 3) showed consistent results when compared to steroids alone (aIRR [95% CI]): IL-6RA (1.13 [0.87,1.45]), combination (0.64 [0.34,1.23]) [Figure 2A]. Linear regression analysis was performed stratified by steroid dose and showed no significant differences between patients received low and high steroid doses compared to IL-6RA (ß = 0.62, 95% CI −1.54,2.78; p=0.57 for low-dose steroid; ß = −1.19, 95% CI −3.85,1.47; p=0.38 for high-dose steroid). Factors associated with higher odds of VFD score were non-use of paralytics, the use of therapeutic anticoagulation, and low FiO2 (Table 3).

**Figure 2.**
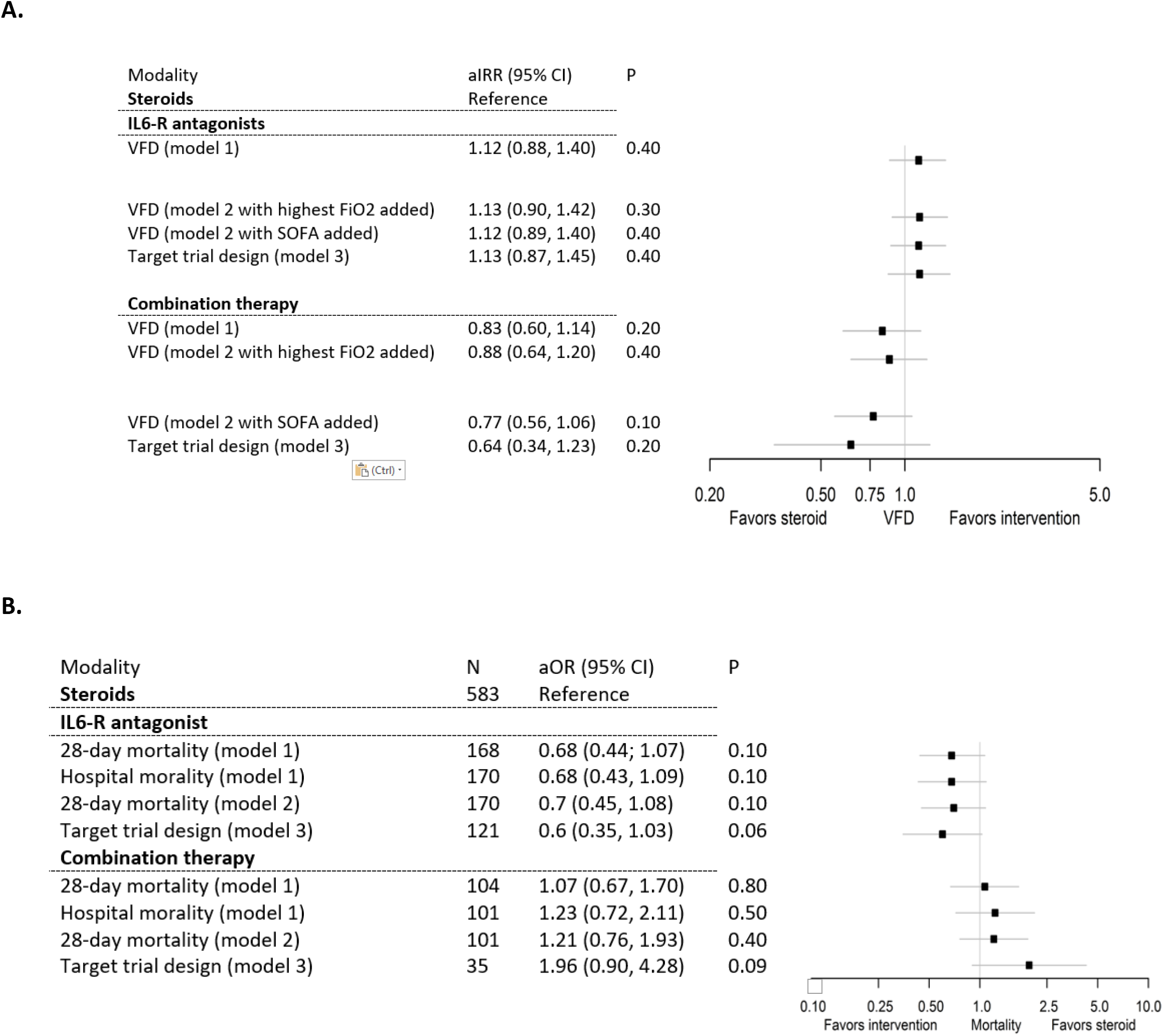
Association between treatment modality, VFD score, and 28-day mortality. (A) Association between treatment modality and ventilator-free days. (B) Association between treatment modality and 28-day mortality. Missing data outcomes in sensitivity analysis (model 3): 4 patients had 28-day mortality status missing and 15 patients had duration of iMV missing. aIRR= adjusted incident rate ratio, aOR= adjusted odds ratio, FiO2 = fractional inspired oxygen concentration, SOFA= Sequential Organ Failure Assessment; VFD = ventilation free days score

**Table 3.** Linear regression analysis for ventilation-free days after adjusting for covariates stratified by steroid dose

| *Predictors* | $\beta$ | 95% CI | *P* |
| --- | --- | --- | --- |
| **Group** |  |  |  |
| **IL-6 antagonist** | *Reference* |  |  |
| **Steroid Low-dose** | 0.62 | -1.54 – 2.78 | 0.57 |
| **Steroid High-dose** | -1.19 | -3.85 – 1.47 | 0.38 |
| Age | -0.01 | -0.07 – 0.04 | 0.59 |
| Weight | -0.02 | -0.05 – 0.00 | 0.07 |
| Lowest FiO2 | 3.20 | 1.26 – 8.27 | **0.02** |
| **Gender** |  |  |  |
| Female | *Reference* |  |  |
| Male | -1.09 | -2.58 – 0.40 | 0.15 |
| **Neuromuscular blocker** |  |  |  |
| Yes | *Reference* |  |  |
| No | 2.87 | 0.18 – 5.55 | **0.04** |
| **Vasopressors** |  |  |  |
| Yes | *Reference* |  |  |
| No | -1.69 | -4.17 – 0.79 | 0.18 |
| **Therapeutic anticoagulation** |  |  |  |
| No | *Reference* |  |  |
| Yes | 2.02 | 0.26 – 3.78 | **0.03** |
| **Any anticoagulation** |  |  |  |
| Yes | *Reference* |  |  |
| No | -1.36 | -3.45 – 0.73 | 0.20 |
| **Ethnicity** |  |  |  |
| Hispanic | *Reference* |  |  |
| Non-Hispanic | 0.67 | -1.20 – 2.54 | 0.48 |
| **Hydroxychloroquine** |  |  |  |
| Yes | *Reference* |  |  |
| No | 1.36 | -0.57 – 3.30 | 0.17 |
| **Antivirals** |  |  |  |
| Yes | *Reference* |  |  |
| No | 0.04 | -1.87 – 1.95 | 0.97 |
| **Azithromycin** |  |  |  |
| Yes | *Reference* |  |  |
| No | -0.28 | -1.85 – 1.30 | 0.73 |
| **ARDS grade** |  |  |  |
| No | *Reference* |  |  |
| Mild (P:F 200-300) | -3.47 | -7.78 – 0.84 | 0.11 |
| Moderate (P:F 100-199) | -1.27 | -3.96 – 1.41 | 0.35 |
| Severe (P:F< 100) | -1.63 | -3.88 – 0.62 | 0.16 |
| **CVD** |  |  |  |
| No | *Reference* |  |  |
| Yes | -0.97 | -2.79 – 0.85 | 0.29 |
| **Asthma/COPD** |  |  |  |
| No | *Reference* |  |  |
| Yes | 0.40 | -1.45 – 2.25 | 0.67 |

The normality of residuals was inspected to ensure good model fit for the linear regression model.
ARDS = Acute respiratory distress syndrome; CI= confidence interval; CVD = cardiovascular diseases; COPD = Chronic obstructive pulmonary disease; P:F = ratio of arterial oxygen partial pressure (PaO2 in mmHg) to fractional inspired oxygen (FiO2); $\beta$ = the log of incident rate ratio. If 0, then it is not significant

### Secondary Outcomes

There was no significant difference in 28-day mortality between IL-6RA alone or in combination compared to steroids alone with different steroids regimen in model 1 (aOR [95% CI]): IL-6RA (0.68 [0.44,1.07]), combination (1.07 [0.67,1.70]). Similarly, exploratory analyses (model 2), sensitivity analysis findings as target trial approach (model 3), and hospital mortality were consistent with primary analysis when compared to standalone steroids (hospital mortality aOR [95% CI]: IL6-RA (0.68, [0.43,1.09]), combination (1.23, [0.72,2.11]) [Figure 2B]. Cause-specific Cox proportional hazard regression showed that the hazard of mortality was not significantly different between IL-6RA (HR = 0.9, p>0.05) or combination (HR = 0.84, p>0.05) compared to steroids (Figure 3). Detailed analysis of secondary outcomes is available in Supplemental Tables 13-17, Figures 1-3. Binary logistic regression was used to assess 28-day-mortality stratified by steroid dose and showed no significant association between use of steroids (low-or high-dose) and mortality compared to IL-6RA (alone or combined). Weighted Kaplan-Maier estimates showed no statistically significant differences between the groups for ICU and hospital LOS (Log-rank p=0.52 and 0.18, respectively). Changes in PF ratio, and incidence of bacteremia were not significantly different as well. IL-6RA patients were more likely to experience liver dysfunction (23.9%) than steroids (14.6 %) or combination (11.9%) (p=0.04) while high troponin levels were reported in combination therapy.

**Figure 3.**
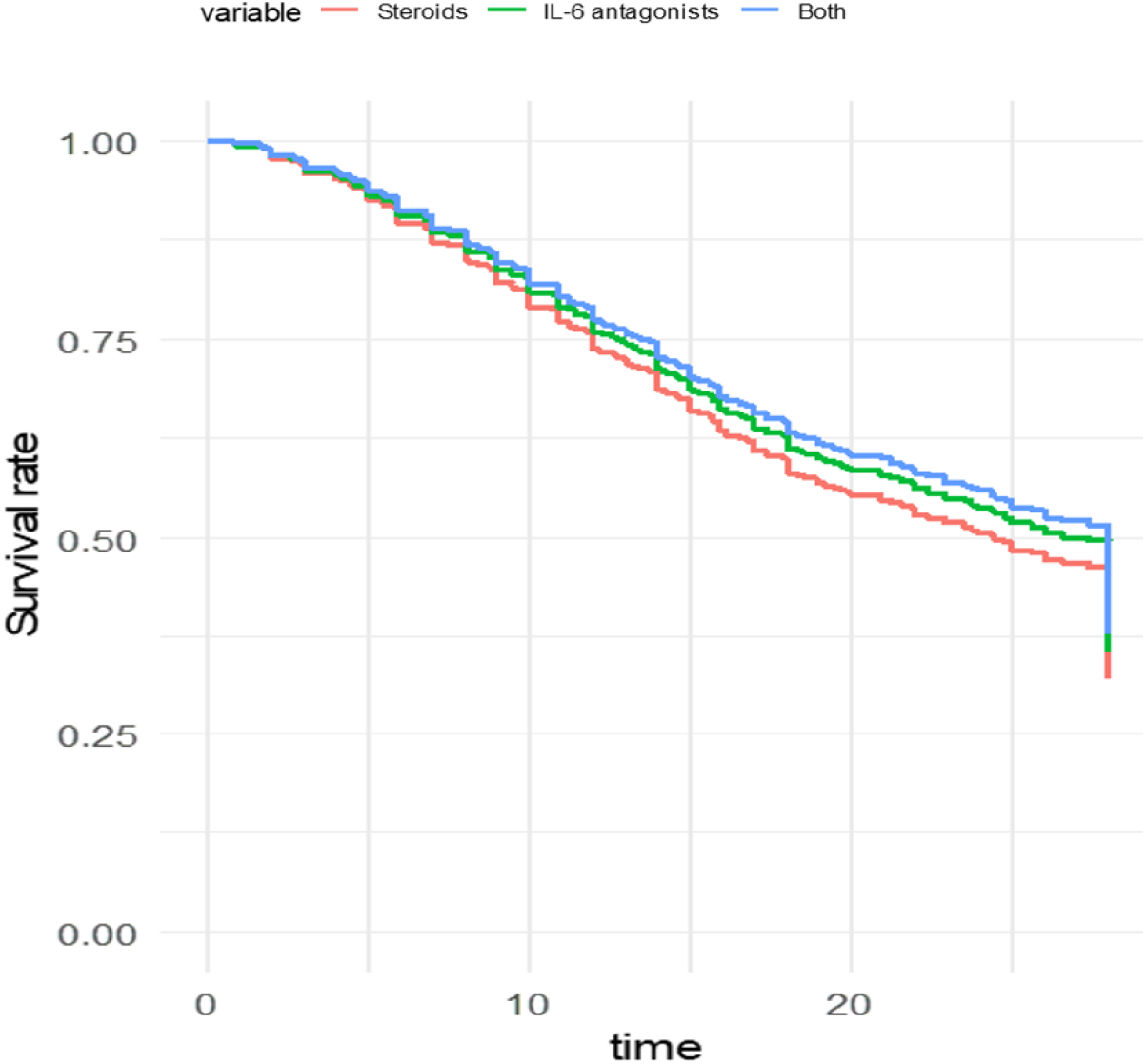
Adjusted survival curves for competing risk models for mortality. Risk regression models for survival endpoints in the presence of competing risks were fitted using binomial regression based on a time sequence of binary event status variables. The same predictors used for marginal structured model were used in the cause-specific Cox regression analysis.

## Discussion

The IL-6RAs were previously investigated with conflicting results (24). Some trials found lower duration of ICU and hospital stays, lower MV and death composite rate, and increased number of organ support-free days in COVID-19 patients requiring respiratory support (8,9,25–29). A recent meta-analysis, trial sequential analysis (TSA), and meta-regression of RCTs by Snow TAC et al showed that the use of tocilizumab may be associated with a short-term mortality benefit in patients with COVID-19: OR 0.87 [0.74–1.01]; p=0.07; TSA adjusted CI 0.66–1.14 (30). There is an ongoing need exists to better identify subgroups of patients who will benefit the most from immunomodulation therapy according to the degree of respiratory support. Herein, we reported characteristics and outcomes of adult ICU COVID-19 patients, who required iMV, and received IL-6RA or steroids within large, multinational registry. We observed no significant difference in outcomes among IL6RA, steroids, and combination therapy and among those who received IL-6R antagonists compared to low-and high-dose steroids.

We proposed several reasons may explain our findings. First, the information on IL6-RA monotherapy use was collected prior to June, 2020 before announcement of RECOVERY steroids domain results. Before this time, we observed steroids use was limited due to concern of delayed viral clearance as shown in MERS coronavirus (31). This may explain the findings and point estimates that were seen here with standalone IL6-RA arm. Whether the effect of IL6-RA monotherapy is beneficial remains unclear and unlikely to be investigated in adequately powered RCT because the equipoise for withholding corticosteroids is no longer present given the convincing evidence for steroids which became standard care for ICU COVID-19 patients requiring MV (32).

Second, and most importantly, our study included only patients with iMV while RECOVERY or REMAP-CAP included varying degrees of respiratory support (iMV at baseline constitute 29 % in REMAP-CAP, and 14 % in RECOVERY) [8,9]. Therefore, the beneficial effect observed in RECOVERY or REMAP-CAP with combined IL6-RA and steroid therapy may be due to patients’ enrollment early in their critical care trajectory while the patients in our study at more severe end of the spectrum. This is in line with the recent Bayesian reanalysis of RECOVERY trial which showed COVID-19 hospitalized patients on non-invasive ventilation have a high probability of a clinically meaningful outcome benefit from tocilizumab and uncertainty for its effect in patients on iMV (15). Taken all together, we think the immunomodulatory effect of IL6-RA and steroid appears to be most beneficial for patients on high-flow nasal cannula, or non-invasive ventilation soon after the time of clinical deterioration at systemic hyperinflammation onset and before intubation or multi-system organ failure (14, 15).

Third, different types and doses of steroids including high-dose steroids in our analysis which could have muted the beneficial effect that IL6-RA might otherwise have had. The REMAP-CAP included fixed duration steroids and shock dependent hydrocortisone and RECOVERY used low-dose dexamethasone 6 mg IV daily. The Italian National Institute for Infectious Diseases recommends 10-day regimen: 5-day of methylprednisolone 1 mg/kg or dexamethasone 20 mg daily (defined as high-dose steroids in our study) (33). This regimen is higher than fixed dexamethasone dose in RECOVERY and that described in non-COVID ARDS literature (12, 13). It was also noted in 19.2% of our study cohort. Prior studies suggested moderate-to-high dose steroid regimens resulted in greater reduction in mortality, organ dysfunction, MV requirement, and no increase in medical or infectious complications compared to low-dose regimens in ICU COVID-19 patients (34, 35). Whether the beneficial immunomodulatory effects of IL-6R blockade could be achieved more easily and with less cost by using a different steroid regimen (higher dose) and its safety need to be investigated in adequately powered RCT (14). Currently, there is an ongoing trial for higher vs. lower doses of dexamethasone for COVID-19 and severe hypoxia (COVIDSTEROID2, NCT04509973) which will include IL6-RA subgroup analysis to provide more information on whether the dose of steroid will have an impact on clinical outcomes (36).

Lastly, racial and ethnic differences in the populations studied, and differences in the study conduction time may provide additional explanation given that the decline in in-hospital mortality from Covid-19 occurred in later phase of the pandemic (24).

Strengths of this study lie in multinational nature. We used data from the largest COVID-19 registry and represented spectrum of intensive care with racially and ethnically diverse cohorts, thereby maximizing generalizability. Moreover, we included large numbers of patients on iMV at baseline which was a limited subset of the population included in the previous trials. Additionally, we included laboratory-confirmed COVID-19 PCR tests, thereby minimizing selection or surveillance bias at each center. Lastly, prior observational studies could be biased by immortal time and indication bias, leading to possible overestimation of positive treatment effect (physicians were more likely to use IL6RA in patients who were more likely to survive or sicker as a last-resort salvage attempt) (14, 38). We used a novel statistical analysis to overcome these limitations and application of target trial design which is likely better approach to reduce immortal time bias and best resembling clinical practice (21–23). Although COVID-19 treatments were not used uniformly before matching and patients did not receive up-to-date standards of care, especially in early pandemic phases, we utilized rigorous methods to match the three groups concerning anticoagulation use, hydroxychloroquine, antivirals, and azithromycin. Importantly, this study reinforces risk of liver injury with IL-6 inhibition. In REMAP-CAP, safety assessment was limited by low AEs number. Clinicians should be aware of these effects and should weigh treatment risks and benefits. Whether this risk is intensified with repeated IL-6RA administration or related to patients’ critical presentation warrants investigation in future studies.

We acknowledge several limitations. First, due to the observational nature of this study, these results should be understood as possibly inconclusive due to insufficient power and frequentist statistical framework. Therefore, there is a possibility of type II errors. Adequately powered RCTs are still needed to further explore the causal treatment effect and clinical outcomes in ICU patients with iMV. Second, fair number of patients were excluded due to missing outcomes and incomplete data for time-dependent confounders to be used in MSM. It is possible that we may have missed patients that would have met eligibility criteria but were not included because of those reasons (possible elimination for non-random missingness). Although we used best available methods to compare well-balanced groups, controlling for confounders in observational study may remain incomplete despite all efforts. Notably, some imbalances in potential confounders (e.g., sex, cardiovascular disease, treatments offered) were observed after weighting. Such potential residual confounding may have a role in the observed non-difference between treatment strategies. Moreover, there was large proportion of incomplete data for SOFA and APACHE II scores, because of heavy burden experienced by participating clinicians during pandemic. We also did not report steroid and IL-6RA administration time from symptom onset and did not address potential role of IL-6 level in predicting IL-6RA responses, all-important questions warranting further investigation. Steroid duration was also highly variable with inconsistent data entry, making it impossible to fully minimize its effect. Finally, while our follow-up is limited to 28 days, additional follow-up may help characterize long-term sequelae, especially mucormycosis, aspergillosis, pneumocystis pneumonia, and multi-drug resistant organisms. Future adequately powered RCT should focus on assessing the survival as primary endpoint which is important to reflect patients, family members, clinicians, and regulators perspectives. Treating steroid dose as a continuous variable and its impact on 28-day mortality will be explored in the subsequent iterations of the registry through ancillary studies by conducting a meta-regression analysis. Going forward, we propose to include research findings, including ours, into the VIRUS web-based decision aid to evaluate risk, benefit, and burden of specific interventions considering prior knowledge, clinical context, preference, and how aforementioned parameters are modified by new findings.

## Conclusions

In adult ICU COVID-19 patients on iMV, we observed no significant difference in outcomes between IL-6RA, steroids, or combination therapy. The lessons learned from this study may suggest a one-size-fits-all for the immunomodulatory effect of IL6-RA and steroid therapy is unlikely to be the answer. Their effect appears to be less pronounced if administered for subsets of patients with iMV. While observational studies cannot suggest causality, further RCTs are needed to enhance the clinicians’ personalized clinical decision-making for COVID-19-associated ARDS.

## Data Availability

The datasets used and analyzed during the current report are available from the corresponding
author on reasonable request.

## Acknowledgments

We thank the SCCM Discovery VIRUS data registry and the Collaborative Co-authors listed in **Supplementary file 2** for providing and maintaining the database of hospitalized COVID-19 patients which made this study possible. We are particularly grateful to Professor Yaseen M Arabi at King Saud bin Abdulaziz University for his inputs and review of this manuscript. We thank the ICU physicians, nurses, respiratory therapists, physiotherapists, pharmacists across the globe for their efforts to save the lives of ICU patients and their braveness in fighting the COVID-19 pandemic.

## Author contributions

MA: proposed the study, led the manuscript writing and did the data collection. AK: made the statistical analysis of this study. All authors made substantial contributions to the concept, design, data collection, and have further participated in the analysis and interpretation of the data. All authors participated in revising the manuscript for important intellectual content and approved the final version to be published. We confirm that the authorship followed the uniform requirements for manuscripts submitted to biomedical journals.

## Availability of data and material

The datasets used and analyzed during the current report are available from the corresponding author on reasonable request.

## Supplemental Content

**Supplemental Table 1.** Included Hospitals

| **Country** | **N** |
| --- | --- |
| 1. United States | 60 |
| 2. Japan | 4 |
| 3. Saudi Arabia | 4 |
| 4. India | 2 |
| 5. Belgium | 2 |
| 6. Colombia | 1 |
| 7. Mexico | 1 |
| 8. Bosnia and Herzegovina | 1 |
| 9. Croatia | 1 |
| 10. Egypt | 1 |
| 11. Hungary | 1 |
| 12. Honduras | 1 |
| 13. Russia | 1 |
| 14. Serbia | 2 |
| 15. Spain | 1 |
| 16. Pakistan | 1 |
| **Total** | **84** |

These are the included hospital per country in the VIRUS registry

**Supplementary Table 2.** Definitions of Selected Variables and Clinical Outcomes

|  |  |
| --- | --- |
| Severity of illness was evaluated using Sequential Organ Failure Assessment (SOFA) and Acute Physiology and Chronic Health Evaluation (APACHEII) scores. | The worst value for baseline laboratory tests was chosen. These data were collected within the first 24 h after admission to the ICU. If that data were not available, then data closest to admission were recorded. |
| Duration of mechanical ventilation | Recorded as the number of calendar days from intubation to extubation or until ICU discharge, or death whichever occurs first. The median ventilator-free days was calculated as calendar days with no ventilator support to day 28. Participants who die before day 28 are assigned zero free days. |
| Secondary infections | Identified based on positive blood, urine, and sputum cultures. Only infections occurring after administration of interleukin-6 Receptor Antagonists or steroids were considered. The definition of infection outcomes was following VIRUS Standard operating definitions |
| 28-day mortality | Defined as the time from hospital admission until death within 28 days, with censoring of alive patients at 28 days. |
| Acute respiratory distress syndrome (ARDS) | Classified according to the Berlin definition [1].<br>The PF ratio was calculated by dividing the arterial Po<sub>2</sub> by the FiO<sub>2</sub> at the same time. If the FiO<sub>2</sub> was not available, the lowest FiO<sub>2</sub> was used followed by the highest FiO<sub>2</sub>. |
| Critical COVID-19 | Defined by the WHO criteria for ARDS, sepsis, septic shock or other conditions that would normally require the provision of life-sustaining therapies, such as mechanical ventilation (invasive or non-invasive) or vasopressor therapy [2]. |
| ICU and hospital length of stay | the interval between the ICU or hospital admission date to the ICU or hospital discharge date was calculated in days |

### Supplementary file 1. Additional details for statistical analysis

International System of Units (SI) were used to represent laboratory data unless otherwise specified. The data were checked for integrity and outliers in the laboratory data (numbers > the 99th percentile) were removed based on clinical judgment and whether the extreme values were clinically plausible.

#### Marginal structural modeling (MSM)

allows for proper adjustment of time-dependent confounding variables, immortal time bias, and indication bias. Immortal time bias refers to the requirement that patients survive long enough to receive the intervention of interest, leading to a potential incorrect overestimation of a positive treatment effect. Indication bias from time-varying confounding variables refers to having an association related to the indication of the intervention that evolves throughout the course of an illness.

MSM using propensity scores were used to compare the primary and secondary outcomes between the three regimens after adjusting for non-time and time-varying covariates [1]. All patients had baseline data on ICU admission. However, the follow-up time varied between patients. Moreover, some variables, such as therapeutic anticoagulation, varied daily. MSMs were used to adjust for variability between patients who received the three regimens. Thus, handling time-varying covariates when weighing observations may have the following limitations:

1. the longitudinal outcome and time-varying covariate may not be available at the same time points: 2. the longitudinal outcome at a particular time point, t, may depend not only on the value of the covariate at the same time point but also at those at other time points. Thus, weighted regressions may be used to account for time-varying confounders using MSMs.

For dichotomous outcomes, the corresponding mixed effects model, namely a mixed effects logistic regression gives fixed effects coefficients that have an interpretation conditional on the random effects. Most often, this is not the interpretation we want. The Generalized Estimating Equations (GEE) approach does give coefficients with a marginal / population-averaged interpretation.

However, an additional practical point that also need to consider is missing data and almost always we have to deal with incomplete data. With regard to this point, mixed models give valid results under the less stringent missing at random assumption compared to the (standard not weighted) GEE that give valid results under the less realistic missing completely at random assumption.

Taking both points (i.e., interpretation and missing data) into account, would most often like to fit a mixed model to be more protected for the missing data but want to obtain parameters that have a population averaged interpretation. An early solution towards this direction was the marginalized mixed models proposed by Heagerty (2), but, in general, these are computationally intensive to fit. A more recent approach that seems to solve the problem has been proposed by Hedeker et al (3). This is implemented in the function marginal_coefs() of the R package GLMMadaptive.

The fixed effects estimates in mixed models with nonlinear link functions have an interpretation conditional on the random effects. However, often we wish to obtain parameters with a marginal / population averaged interpretation, which leads many researchers to use generalized estimating equations, and dealing with potential issues with missing data. Nonetheless, recently Hedeker et al. have proposed a nice solution to this problem. Their approach is implemented in function marginal_coefs(). For example, for model fm we obtain the marginalized coefficients.

The function calculates the marginal log odds ratios in our case (because we have a binary outcome) using a Monte Carlo procedure with number of samples determined by the M argument. Standard errors for the marginalized coefficients are obtained by setting std_errors = TRUE in the call to marginal_coefs(), and require a double Monte Carlo procedure for which argument K comes also into play.

Generalized estimating equations (GEE) were constructed to evaluate the impact of the treatment (t) on the outcome (d) and to handle the time-varying covariate (v) in the panel dataset. Since the outcome was a continuous variable, a generalized linear Gaussian family with identity link was used. Auto-regressive (AR1) correlation structure was selected since this was a time series (panel) data; we expected the correlation to decay as the outcome values were farther away from the time of interest.

**Covariates included in the model were age, sex, ethnicity, asthma/COPD, ARDS grade (none, mild, moderate or severe) at admission, history of CVD (diabetes, hypertension, coronary heart diseases or congestive heart failure), and the baseline lowest FiO2. Other covariates included the use of any anticoagulation, therapeutic anticoagulation, hydroxychloroquine, azithromycin, and antivirals, (including remdesivir); vasopressors; and neuromuscular blockers**. The effect of adding steroids, IL-6 antagonists was assessed by including each variables as a dummy coded variable in the marginal model. In addition, an interaction term was added to the model to assess whether the combination of both drugs was associated with a statistically significant effect size.

inverse probability weighting (IPTW) are weights assigned to each observation across time conditioned on the previous exposure history, which are then multiplied to generate a single weight for a subject (4). Similar to conventional propensity score estimation, IPTW is generated using either a logit or probit model that regresses covariates to a treatment group (exposure) variable. With IPTW, the previous exposure history is incorporated to the propensity score estimation, which is time-varying. Standardized weights in a longitudinal setting are estimated. The numerator contains the probability of the observed exposure at each time point conditioned on the observed exposure history of the previous time point and the observed non-time varying covariates. The denominator contains the probability of the observed exposure at each time point conditioned on the observed exposure history of the previous time point, the observed time-varying covariates history at the current time point, and the non-time varying covariates. In standardized weights, the time-varying confounders are captured in the denominator but not in the numerator. However, the non-time varying (also known as fixed-time) covariates are captured in both the numerator. Due to the longitudinal nature of the data, the GLMMadaptive package was used for model fitting. The Package GLMMadaptive provides a suit of functions for fitting and post-processing mixed effects models for grouped/clustered outcomes which have a distribution other than a normal distribution. In particular, let yi denote a vector of grouped/clustered outcome for the i-th sample unit (i=1,…,n).

Standardized mean difference (SMD) were reported for comparison between cohorts before and after matching

#### Handling of missing data

The generalized boosted model (GMB) creates a separate level during matching called NA and creates weights based on the distribution of missing values. So basically it considers NA as a level of its own. The weights are created for all respondents and are taking the missing values into consideration. Imputation requires that data is missing at random which is not the case here. Moreover, the variables we used had few missing values so there is no problem which would not have been the case if we used the SOFA and APACHE which had a lot of missing data.

#### The pseudo population

The populations are weighed by the probability of receiving treatments based on other confounders. The weights were produced using generalized boosted models and these weights were used when doing further analysis. The pseudo population is the sum of weights for each group rather than considering each observation as 1. Thus, the numbers can increase or decrease compared to the initial number of the group depends on the assigned weights. The aim to emulate randomized trial to create a pseudo study cohort, where the weighted version can balance off the covariate bias and mimic a randomized treatment assignment situation

---

**Supplementary Table 3.** Distribution of baseline covariates across groups before adjustment

|  | **Steroids** | **IL6-R antagonists** | **Both** | **P** |
| --- | --- | --- | --- | --- |
|  | **N=589** | **N=170** | **N=101** |  |
| **Sex** |  |  |  | 0.39 |
| Female | 205 (34.8) | 49 (28.8) | 34 (33.7) |  |
| Male | 384 (65.2) | 119 (70) | 70 (66.3) |  |
| **Age, mean (SD)** | 61.1 (14.3) | 57.4 (14.7) | 62.1 (13.2) | 0.01 |
| **Therapeutic anticoagulation** |  |  |  | 0.24 |
| No | 455 (77.2) | 136 (80) | 75 (74.3) |  |
| Yes | 134 (22.8) | 32 (20) | 29 (25.7) |  |
| **Ethnic group** |  |  |  | <0.001 |
| Hispanic | 112 (19) | 48 (28.2) | 22 (21.7) |  |
| Non-Hispanic | 263 (44.7) | 80 (47.1) | 55 (54.4) |  |
| **Weight (kg) at Hospital Admission, mean (SD)** | 90.1 (27.7) | 96.9 (28) | 95.3 (28.5) | 0.01 |
| **Cardiovascular disease** | 130 (22.1) | 16 (9.4) | 23 (22.7) | 0.001 |
| **Asthma/COPD** | 102 (17.3) | 19 (11.1) | 29 (28.7) | 0.002 |
| **Hydroxychloroquine** | 87 (14.8) | 87 (51.2) | 41 (40.6) | <0.001 |
| **Antiviral** | 474 (80.5) | 159 (93.5) | 86 (85.1) | <0.001 |
| **Azithromycin** | 204 (34.6) | 93 (54.7) | 50 (49.5) | <0.001 |
| **ARDS** |  |  |  | . |
| No | 380 (67.3) | 144 (84.7) | 67 (66.3) |  |
| Mild (P:F 200-300) | 15 (2.7) | 4 (2.4) | 6 (5.9) |  |
| Moderate (P:F 100-199) | 65 (11.5) | 4 (2.4) | 8 (7.9) |  |
| Severe (P:F< 100) | 105 (18.6) | 12 (7.1) | 16 (15.8) |  |
| **Lowest FiO2, mean (SD)** | 0.57 (0.3) | 0.51 (0.3) | 0.55 (0.2) | 0.12 |

Data presented as n (%) unless otherwise specified
Abbreviation: ARDS: Acute respiratory distress syndrome ; COPD: Chronic obstructive pulmonary disease SD: standard deviation; P:F: ratio of arterial oxygen partial pressure (PaO2 in mmHg) to fractional inspired oxygen (FiO2)

**Supplementary Table 4.** Other Baseline characteristics of the total cohort

|  | **Steroids**<br>**N=589** | **IL6-R**<br>**antagonists**<br>**N=170** | **Both**<br>**N=101** | **P** | **N** |
| --- | --- | --- | --- | --- | --- |
| **Hospital Admission Source** |  |  |  | <0.001 | 858 |
| Home | 207 (35.3) | 42 (24.7) | 36 (35.6) |  |  |
| Hospital ED | 177 (30.2) | 92 (54.1) | 43 (42.6) |  |  |
| Nursing Home | 25 (4.26) | 2 (1.2) | 2 (1.9) |  |  |
| Other | 5 (0.85) | 1 (0.6) | 0 (0) |  |  |
| Outside ED | 30 (5.11) | 8 (4.7) | 3 (2.9) |  |  |
| Transfer from other Facility | 143 (24.4) | 25 (14.7) | 17 (16.8) |  |  |
| **Signs and symptoms** |  |  |  | 0.80 | 860 |
| Abdominal pain | 38 (6.5) | 5 (2.9) | 7 (6.9) | 0.20 | 860 |
| Ageusia (loss of taste sense) | 25 (4.24) | 5 (2.9) | 3 (2.9) | 0.82 | 860 |
| Loss of appetite | 51 (8.7) | 25 (14.7) | 13 (12.9) | 0.05 | 860 |
| Anosmia (loss of smell sense) | 19 (3.2) | 4 (2.4) | 6 (5.9) | 0.27 | 860 |
| Arthralgia | 11 (1.9) | 4 (2.4) | 6 (5.9) | 0.05 | 860 |
| Chest pain/tightness | 67 (11.4) | 16 (9.4) | 11 (10.9) | 0.77 | 860 |
| Chills/rigors | 84 (14.3) | 33 (19.4) | 15 (14.9) | 0.26 | 860 |
| Confusion/delirium | 49 (8.3) | 11 (6.5) | 7 (6.9) | 0.69 | 860 |
| Conjunctival congestion | 1 (0.17) | 1 (0.59) | 0 (0) | 0.53 | 860 |
| Dry cough | 283 (48) | 101 (59.4) | 58 (57.4) | 0.01 | 860 |
| Cough with sputum | 69 (11.7) | 25 (14.7) | 16 (15.8) | 0.37 | 860 |
| Diarrhea | 78 (13.2) | 44 (25.9) | 21 (20.8) | **<0.001** | 860 |
| Dizziness/lightheadedness | 31 (5.3) | 8 (4.7) | 5 (4.9) | 0.96 | 860 |
| Dyspnea/shortness of breath | 436 (74) | 133 (78.2) | 73 (72.3) | 0.45 | 860 |
| Fever | 370 (62.8) | 125 (73.5) | 72 (71.3) | **0.02** | 860 |
| Headache | 60 (10.2) | 23 (13.5) | 11 (10.9) | 0.47 | 860 |
| Hemoptysis | 7 (1.2) | 0 (0) | 0 (0) | 0.39 | 860 |
| Malaise | 91 (15.4) | 28 (16.5) | 23 (22.8) | 0.19 | 860 |
| Myalgia or fatigue | 439 (74.5) | 101 (59.4) | 62 (61.4) | **<0.001** | 860 |
| Nasal congestion/rhinorrhea | 25 (4.2) | 12 (7.06) | 10 (9.9) | **0.04** | 860 |
| Nausea and Vomiting | 81 (13.8) | 22 (12.9) | 15 (14.9) | 0.91 | 860 |
| Night sweat | 9 (1.5) | 1 (0.59) | 2 (1.9) | 0.60 | 860 |
| Seizure | 6 (1.02) | 1 (0.59) | 0 (0) | 0.85 | 860 |
| Sneezing | 1 (0.2) | 1 (0.59) | 0 (0) | 0.53 | 860 |
| Sore throat or throat pain | 33 (5.6) | 15 (8.8) | 11 (10.9) | 0.08 | 860 |
| Swollen neck/glands (lymphadenopathy) | 1 (0.2) | 0 (0) | 0 (0) | 1.00 | 860 |
| **Comorbidities** |  |  |  |  |  |
| Coronary heart diseases | 492 (83.5) | 161 (94.7) | 85 (84.2) | **0.001** | 860 |
| Hypertension | 251 (42.6) | 67 (39.4) | 45 (44.6) | 0.67 | 860 |
| Cardiac arrhythmias | 50 (8.5) | 5 (2.9) | 4 (3.9) | **0.02** | 860 |
| Congestive heart failure | 72 (12.2) | 10 (5.9) | 9 (8.9) | **0.05** | 860 |
| Valvular heart disease | 17 (2.9) | 1 (0.6) | 1 (0.9) | 0.18 | 860 |
| Chronic pulmonary disease not asthma | 59 (10) | 7 (4.1) | 16 (15.8) | **0.005** | 860 |
| Asthma physician diagnosed | 45 (7.6) | 12 (7.1) | 12 (11.9) | 0.31 | 860 |
| Pulmonary circulation disorder | 7 (1.2) | 1 (0.6) | 2 (1.9) | 0.55 | 860 |
| Chronic kidney disease | 58 (9.9) | 14 (8.24) | 12 (11.9) | 0.62 | 860 |
| Chronic dialysis | 12 (2.04) | 2 (1.18) | 3 (2.9) | 0.59 | 860 |
| Diabetes | 362 (61.5) | 111 (65.3) | 55 (54.5) | 0.21 | 860 |
| Hypothyroidism | 45 (7.6) | 6 (3.5) | 5 (4.9) | 0.13 | 860 |
| Liver disease | 20 (3.4) | 2 (1.2) | 2 (1.9) | 0.32 | 860 |
| Hepatitis B | 2 (0.34) | 1 (0.6) | 0 (0) | 0.68 | 860 |
| Hepatitis C | 11 (1.9) | 3 (1.8) | 2 (1.9) | 1.00 | 860 |
| peptic ulcer disease excluding bleeding | 6 (1.02) | 3 (1.8) | 2 (1.9) | 0.44 | 860 |
| solid tumor without metastasis | 26 (4.4) | 0 (0) | 8 (7.9) | **0.001** | 860 |
| hematologic malignancy | 7 (1.2) | 4 (2.4) | 1 (0.9) | 0.43 | 860 |
| metastatic cancer | 10 (1.7) | 0 (0) | 0 (0) | 0.12 | 860 |
| history of solid organ or bone marrow | 1 (0.17) | 0 (0) | 2 (1.9) | 0.06 | 860 |
| HIV aids or other immunosuppression | 6 (1.02) | 4 (2.4) | 2 (1.9) | 0.24 | 860 |
| stroke or other neurological disorders | 47 (7.9) | 4 (2.4) | 5 (4.9) | **0.03** | 860 |
| Paralysis | 5 (0.9) | 1 (0.6) | 0 (0) | 1.00 | 860 |
| Rheumatoid arthritis collagen vascular disease | 9 (1.5) | 3 (1.8) | 1 (0.9) | 0.91 | 860 |
| Blood loss anemia | 0 (0) | 1 (0.6) | 0 (0) | 0.32 | 860 |
| Iron deficiency anemia | 13 (2.2) | 3 (1.8) | 2 (1.9) | 1.00 | 860 |
| Coagulopathy | 4 (0.68) | 3 (1.8) | 1 (0.9) | 0.32 | 860 |
| Malnutrition | 1 (0.17) | 0 (0) | 1 (0.9) | 0.26 | 860 |
| Obesity | 117 (19.9) | 60 (35.3) | 32 (31.7) | **<0.001** | 860 |
| Substance use disorder | 15 (2.6) | 2 (1.2) | 2 (1.9) | 0.63 | 860 |
| Depression | 37 (6.3) | 5 (2.9) | 10 (9.9) | 0.06 | 860 |
| Psychosis | 5 (0.9) | 1 (0.6) | 0 (0) | 1.00 | 860 |
| Dementia | 18 (3.1) | 3 (1.8) | 2 (1.9) | 0.76 | 860 |
| Obstructive sleep apnea | 33 (5.6) | 19 (11.2) | 11 (10.9) | **0.02** | 860 |
| Venous thromboembolism | 15 (2.6) | 4 (2.4) | 6 (5.9) | 0.19 | 860 |
| Dyslipidemia hyperlipidemia | 59 (10) | 6 (3.5) | 10 (9.9) | **0.03** | 860 |
| **Admission Diagnosis/Complications** |  |  |  |  |  |
| Acute Liver Injury | 22 (3.7) | 5 (2.9) | 6 (5.9) | 0.42 | 860 |
| Acute Myocardial Infarction | 9 (1.5) | 1 (0.6) | 0 (0) | 0.41 | 860 |
| Acute Renal Failure Requiring Hemofiltration | 23 (3.9) | 5 (2.9) | 8 (7.9) | 0.12 | 860 |
| Acute Renal Injury | 89 (15.1) | 31 (18.2) | 14 (13.9) | 0.54 | 860 |
| Bacteremia | 16 (2.7) | 4 (2.4) | 0 (0) | 0.29 | 860 |
| Bacterial Pneumonia | 59 (10) | 23 (13.5) | 8 (7.9) | 0.28 | 860 |
| Cardiac Arrest | 19 (3.2) | 1 (0.6) | 1 (0.9) | 0.09 | 860 |
| Cardiac Arrhythmia: Atrial Fibrillation | 10 (1.7) | 3 (1.8) | 4 (3.9) | 0.35 | 860 |
| Cardiac Arrhythmia: Heart Block | 2 (0.3) | 0 (0) | 1 (0.9) | 0.40 | 860 |
| Cardiac Arrhythmia: Ventricular Tachycardia | 8 (1.4) | 0 (0) | 2 (1.9) | 0.19 | 860 |
| Congestive Heart Failure / Cardiomyopathy | 13 (2.2) | 1 (0.6) | 3 (2.9) | 0.31 | 860 |
| Diabetic Ketoacidosis (DKA) | 2 (0.34) | 0 (0) | 0 (0) | 1.00 | 860 |
| Delirium / Encephalopathy | 48 (8.2) | 4 (2.4) | 3 (2.9) | **0.008** | 860 |
| Gastrointestinal Hemorrhage | 2 (0.3) | 1 (0.6) | 1 (0.9) | 0.38 | 860 |
| Hyperglycemia | 67 (11.4) | 22 (12.9) | 14 (13.9) | 0.71 | 860 |
| Hypoglycemia | 5 (0.9) | 0 (0) | 0 (0) | 0.63 | 860 |
| Meningitis/Encephalitis | 3 (0.5) | 0 (0) | 0 (0) | 1.00 | 860 |
| Myocarditis | 3 (0.5) | 0 (0) | 1 (0.9) | 0.53 | 860 |
| Pneumothorax | 3 (0.5) | 0 (0) | 2 (1.9) | 0.16 | 860 |
| Pleural Effusion | 6 (1.02) | 0 (0) | 1 (0.9) | 0.48 | 860 |
| Rhabdomyolysis / Myositis | 8 (1.4) | 0 (0) | 2 (1.9) | 0.19 | 860 |
| Seizure | 5 (0.9) | 0 (0) | 0 (0) | 0.63 | 860 |
| Shock | 72 (12.2) | 9 (5.3) | 9 (8.9) | **0.03** | 860 |
| Stroke | 8 (1.4) | 0 (0) | 0 (0) | 0.22 | 860 |
| **Pre-Hospital (Home) Medication** |  |  |  |  |  |
| Angiotensin-converting-enzyme Inhibitor | 97 (16.5) | 31 (18.2) | 20 (19.8) | 0.66 | 860 |
| Angiotensin receptor blocker | 68 (11.5) | 22 (12.9) | 13 (12.9) | 0.85 | 860 |
| Antiretroviral | 2 (0.3) | 2 (1.2) | 1 (0.9) | 0.23 | 860 |
| Antibiotics | 54 (9.2) | 11 (6.5) | 7 (6.9) | 0.46 | 860 |
| Anti-Diabetic | 44 (7.5) | 5 (2.9) | 13 (12.9) | **0.009** | 860 |
| Aspirin | 122 (20.7) | 31 (18.2) | 29 (28.7) | 0.11 | 860 |
| Chemotherapy currently or in the last 3 | 12 (2.04) | 1 (0.6) | 2 (1.9) | 0.51 | 860 |
| Immunotherapy | 9 (1.5) | 0 (0) | 3 (2.9) | 0.11 | 860 |
| Inhaled corticosteroids | 54 (9.2) | 13 (7.7) | 13 (12.9) | 0.35 | 860 |
| NSAID/Ibuprofen | 49 (8.3) | 18 (10.6) | 10 (9.9) | 0.62 | 860 |
| Other anti-hypertensive agent (eg., beta blocker, calcium channel blocker, diuretic) | 211 (35.8) | 57 (33.5) | 37 (36.6) | 0.83 | 860 |
| Paracetamol/Acetaminophen | 81 (13.8) | 25 (14.7) | 22 (21.8) | 0.11 | 860 |
| Proton Pump Inhibitors | 76 (12.9) | 27 (15.9) | 20 (19.8) | 0.15 | 860 |
| Statins | 166 (28.2) | 58 (34.1) | 31 (30.7) | 0.32 | 860 |
| Anticoagulants | 46 (7.8) | 9 (5.3) | 10 (9.9) | 0.35 | 860 |
| Hydroxychloroquine (Plaquenil) | 5 (0.9) | 2 (1.2) | 1 (0.9) | 0.87 | 860 |
| **Social History** |  |  |  |  |  |
| Social history (current smoker) | 29 (4.9) | 7 (4.12) | 6 (5.94) | 0.80 | 860 |
| Social history (former smoker) | 119 (20.2) | 29 (17.1) | 25 (24.8) | 0.31 | 860 |
| Social history (vaping) | 0 (0) | 1 (0.59) | 0 (0.00) | 0.32 | 860 |
| Social history (alcohol use disorder) | 26 (4.4) | 6 (3.53) | 5 (4.95) | 0.81 | 860 |
| Social history (substance use disorder) | 14 (2.4) | 3 (1.76) | 1 (0.99) | 0.81 | 860 |
| Social history (unknown/not available) | 37 (6.3) | 5 (2.94) | 4 (3.96) | 0.19 | 860 |
| **Packs per day (PPD) patient smokes** |  |  |  | 0.09 | 49 |
| < ½ PPD (0.5 PPD) | 8 (21.1) | 0 (0) | 2 (33.3) |  |  |
| 1 PPD | 6 (15.8) | 0 (0) | 2 (33.3) |  |  |
| 1½ PPD (1.5 PPD) | 1 (2.6) | 0 (0) | 0 (0) |  |  |
| 2 PPD | 4 (10.5) | 1 (20) | 2 (33.3) |  |  |
| Unknown/Not Available | 19 (50) | 4 (80) | 0 (0) |  |  |
| Does/Did patient uses other tobacco products regularly? (Cigars) | 6 (1.02) | 0 (0) | 1 (0.9) | 0.48 | 860 |
| Does/Did patient uses other tobacco products regularly? (Pipes) | 1 (0.2) | 0 (0) | 1 (0.9) | 0.26 | 860 |
| Does/Did patient uses other tobacco products regularly? (Chewing tobacco) | 2 (0.3) | 0 (0) | 0 (0) | 1.00 | 860 |
| Does/Did patient uses other tobacco products regularly? (No other tobacco) | 9 (1.5) | 1 (0.6) | 3 (2.9) | 0.27 | 860 |
| Does/Did patient uses other tobacco products regularly? (Unknown/ Not available) | 19 (3.2) | 4 (2.4) | 2 (1.9) | 0.86 | 860 |
| **Type of Shock** |  |  |  | 0.49 | 80 |
| Cardiogenic shock | 5 (7.6) | 0 (0) | 0 (0) |  |  |
| Distributive shock | 59 (89.4) | 5 (83.3) | 8 (100) |  |  |
| Hypovolemic shock | 2 (3.03) | 1 (16.7) | 0 (0) |  |  |

Data presented as n (%) unless otherwise specified
Abbreviation: ARDS: Acute respiratory distress syndrome ; P:F: ratio of arterial oxygen partial pressure (PaO<sub>2</sub> in mmHg) to fractional inspired oxygen (FiO<sub>2</sub>)

**Supplementary Table 5.** Ventilator and Radiology Characteristics of the included patients at days 0 and 1 of ICU stay

|  | Day 0 |  |  |  | Day 1 |  |  |  |
| --- | --- | --- | --- | --- | --- | --- | --- | --- |
|  | Steroids | IL6-R antagonists | combination | P | Steroids | IL6-R antagonists | combination | P |
|  | *N=589* | *N=170* | *N=101* |  | *N=568* | *N=170* | *N=94* |  |
| **Performed diagnostics** |  |  |  |  |  |  |  |  |
| None | 31 (5.3) | 7 (4.1) | 7 (6.9) | 0.60 | 232 (40.8) | 49 (28.8) | 33 (35.1) | **0.02** |
| Chest X-ray | 467 (79.3) | 149 (87.6) | 89 (88.1) | **0.01** | 256 (45.1) | 108 (63.5) | 53 (56.4) | **<0.001** |
| CT Chest | 153 (26) | 25 (14.7) | 13 (12.9) | **<0.001** | 27 (4.75) | 6 (3.5) | 5 (5.3) | 0.73 |
| Lung US | 8 (1.4) | 0 (0) | 1 (0.9) | 0.31 | 2 (0.4) | 1 (0.6) | 0 (0) | 0.68 |
| Cardiac Echo | 26 (4.4) | 3 (1.8) | 2 (1.9) | 0.22 | 30 (5.3) | 4 (2.4) | 3 (3.2) | 0.27 |
| **infiltrates present in CXR** | 424 (91.4) | 136 (91.3) | 84 (94.4) | 0.62 | 244 (97.2) | 105 (97.2) | 53 (100) | 0.72 |
| **Interstitial pattern** | 108 (18.3) | 26 (15.3) | 24 (23.8) | 0.22 | 58 (10.2) | 18 (10.6) | 12 (12.8) | 0.76 |
| Opacities | 320 (54.3) | 103 (60.6) | 62 (61.4) | 0.20 | 194 (34.2) | 80 (47.1) | 37 (39.4) | 0.01 |
| Multifocal | 155 (26.3) | 50 (29.4) | 30 (29.7) | 0.62 | 79 (13.9) | 48 (28.2) | 23 (24.5) | **<0.001** |
| Bilateral | 336 (57) | 102 (60) | 63 (62.4) | 0.53 | 203 (35.7) | 90 (52.9) | 39 (41.5) | **<0.001** |
| Pleural effusion | 26 (4.4) | 6 (3.5) | 8 (7.9) | 0.22 | 15 (2.6) | 6 (3.5) | 4 (4.3) | 0.48 |
| **CT chest pattern<sup>¶</sup>** |  |  |  |  |  |  |  |  |
| Ground Glass Opacity (GGO) | 104 (17.7) | 17 (10) | 13 (12.9) | 0.04 | 19 (3.4) | 6 (3.5) | 5 (5.3) | 0.57 |
| Crazy paving | 10 (1.7) | 1 (0.6) | 2 (1.9) | 0.57 | 2 (0.4) | 0 (0) | 1 (1.1) | 0.40 |
| Multifocal | 88 (14.9) | 14 (8.2) | 8 (7.9) | **0.02** | 11 (1.9) | 3 (1.8) | 2 (2.1) | 1 |
| Bilateral | 134 (22.8) | 18 (10.6) | 12 (11.9) | **<0.001** | 18 (3.2) | 6 (3.5) | 4 (4.3) | 0.76 |
| Lymphadenopathy (LAP) | 13 (2.2) | 1 (0.6) | 1 (0.9) | 0.40 | 3 (0.5) | 0 (0) | 0 (0) | 1 |
| Pleural effusion | 17 (2.9) | 2 (1.2) | 0 (0) | 0.12 | 7 (1.2) | 1 (0.6) | 0 (0) | 0.63 |
| Opacity | 49 (8.3) | 7 (4.1) | 5 (4.9) | 0.11 | 6 (1.1) | 0 (0) | 0 (0) | 0.43 |
| Consolidation | 27 (4.6) | 1 (0.6) | 0 (0) | 0.003 | 3 (0.5) | 1 (0.6) | 1 (1.1) | 0.78 |
| Pulmonary nodules | 2 (0.34) | 0 (0) | 0 (0) | 1.00 | 0 (0) | 0 (0) | 0 (0) | 1 |
| **Respiratory Rate- mean (SD)** | 21.0 (24.2) | 29.8 (61.5) | 17.9 (4.9) | 0.12 | 23.9 (36.2) | 34.2 (75.1) | 18.3 (5.6) | 0.07 |
| **Tidal volume (ml) mean (SD)** | 432 (121) | 439 (114) | 444 (97.8) | 0.83 | 432 (113) | 419 (108) | 442 (93.6) | 0.46 |
| **PEEP (cm H2O) mean (SD)** | 11.2 (3.6) | 12 (3.9) | 11.1 (4.3) | 0.36 | 11.8 (6.4) | 12.0 (3.8) | 11.5 (3.1) | 0.88 |
| **Ventilator mode** |  |  |  | **<0.001** |  |  |  | **<0.005** |
| Airway pressure release ventilation (APRV) | 6 (1.9) | 1 (2) | 6 (16.7) |  | 9 (2.5) | 1 (0.9) | 2 (3.7) |  |
| Pressure control (PC) | 49 (15.5) | 4 (8) | 8 (22.2) |  | 42 (11.5) | 5 (4.9) | 12 (22.2) |  |
| Pressure Support (PS) | 4 (1.3) | 1 (2) | 1 (2.8) |  | 4 (1.1) | 1 (0.9) | 1 (1.9) |  |
| Volume control (VC) | 206 (65) | 22 (44) | 16 (44.4) |  | 243 (66.8) | 63 (62.4) | 32 (59.3) |  |
| **Documented ventilator associated pneumonia (VAP)** | 15 (5.2) | 0 (0) | 4 (10.5) | **0.05** | 13 (3.9) | 3 (2.9) | 2 (3.7) | 1 |
| **Documented assessment of spontaneous breathing trial** |  |  |  | 0.32 |  |  |  | 0.46 |
| No | 145 (50.7) | 28 (54.9) | 18 (48.6) |  | 180 (54.9) | 55 (55) | 26 (48.1) |  |
| not indicated | 131 (45.8) | 22 (43.1) | 15 (40.5) |  | 118 (36) | 39 (39) | 26 (48.1) |  |
| Yes | 10 (3.5) | 1 (1.9) | 4 (10.8) |  | 30 (9.2) | 6 (6) | 2 (3.7) |  |

Data presented as n (%) unless otherwise specified
¶ percentage calculated based on available data for patients with computerized tomography (CT) chest scan

**Supplementary table 6:** SOFA and APCHE-II scores at baseline

|  | Steroids | IL-6 antagonists | Both | P | N |
| --- | --- | --- | --- | --- | --- |
|  | *N=446* | *N=105* | *N=73* |  |  |
| **SOFA Respiration:** |  |  |  | . | 622 |
| 0. >400 | 19 (4.3) | 10 (9.5) | 2 (2.8) |  |  |
| 1. < 400 (S:F 221-301), +/- Respiratory support | 25 (5.6) | 9 (8.6) | 6 (8.5) |  |  |
| 2. < 300 (S:F 142-220), +/- Respiratory support | 73 (16.4) | 19 (18.1) | 19 (26.8) |  |  |
| 3. < 200 (S:F 67-141) and Respiratory support | 165 (37) | 46 (43.8) | 26 (36.6) |  |  |
| 4. < 100 (S:F <67) and Respiratory support | 164 (36.8) | 21 (20) | 18 (25.4) |  |  |
| **SOFA coagulation (platelet):** |  |  |  | 0.41 | 582 |
| 0. >150 | 338 (83.5) | 86 (82.7) | 60 (82.2) |  |  |
| 1. < 150 | 47 (11.6) | 11 (10.6) | 13 (17.8) |  |  |
| 2. < 100 | 14 (3.5) | 6 (5.8) | 0 (0) |  |  |
| 3. < 50 | 5 (1.2) | 1 (0.9) | 0 (0) |  |  |
| 4. < 20 | 1 (0.3) | 0 (0) | 0 (0) |  |  |
| **SOFA cardiovascular (vasopressors):** |  |  |  | . | 583 |
| 0. No Hypotension | 233 (57.5) | 60 (57.1) | 40 (54.8) |  |  |
| 1. MAP < 70 mm Hg | 25 (6.2) | 21 (20) | 11 (15.1) |  |  |
| 2. Dopamine ≤ 5 or Dobutamine (any dose) | 1 (0.3) | 0 (0) | 1 (1.4) |  |  |
| 3. Dopamine > 5 or Epi/Norepi ≤0.1 | 70 (17.3) | 11 (10.5) | 11 (15.1) |  |  |
| 4. Dopamine > 15 or Epi/Norepi > 0.1 | 76 (18.8) | 13 (12.4) | 10 (13.7) |  |  |
| **SOFA GCS:** |  |  |  | 0.01 | 582 |
| 0. 15 | 176 (43.5) | 62 (59) | 38 (52.8) |  |  |
| 1. 13-14 | 37 (9.1) | 9 (8.6) | 4 (5.6) |  |  |
| 2. 10-12 | 30 (7.4) | 9 (8.6) | 6 (8.3) |  |  |
| 3. 6-9 | 73 (18) | 18 (17.1) | 16 (22.2) |  |  |
| 4. <6 | 89 (22) | 7 (6.7) | 8 (11.1) |  |  |
| **SOFA Liver (bilirubin):** |  |  |  | 0.16 | 569 |
| 0. < 1.2 | 335 (84.8) | 92 (89.3) | 64 (90.1) |  |  |
| 1. 1.2 -1.9 mg/dl ..... 20-32 micromole/L | 41 (10.4) | 7 (6.8) | 2 (2.8) |  |  |
| 2. 2.0 - 5.9 mg/dl ..... 33-101 micromole /L | 18 (4.6) | 3 (2.9) | 4 (5.6) |  |  |
| 3. 6.0-11.9 mg/dl ..... 102-204 micromole /L | 1 (0.3) | 1 (0.9) | 1 (1.4) |  |  |
| **SOFA Renal (creatinine or urine output):** |  |  |  | . | 583 |
| 0. < 1.2 mg/dl (110μmol/L) | 254 (62.7) | 73 (69.5) | 46 (63) |  |  |
| 1. 1.2 -1.9 mg/dl (110-170μmol/L) | 77 (19) | 21 (20) | 15 (20.5) |  |  |
| 2. 2.0 - 2.4 mg/dl (171-299μmol/L) | 28 (6.9) | 5 (4.8) | 6 (8.2) |  |  |
| 3. 2.5-4.9 mg/dl (300-440μmol/L) or urine output < 500 mL/day | 30 (7.4) | 5 (4.8) | 4 (5.5) |  |  |
| 4. > 5.0 mg/dl (>440μmol/L) or urine output < 200 mL/day | 16 (3.9) | 1 (0.9) | 2 (2.7) |  |  |
| **Calculated APACHE II (First 24 hours in ICU that is Day 1 ICU)** |  |  |  | <0.001 | 117 |
| No | 23 (46.9) | 4 (9.1) | 6 (25) |  |  |
| Yes | 26 (53.1) | 40 (90.9) | 18 (75) |  |  |
| APACHE II (First 24 hours) score, mean (SD) | 18.1 (7.1) | 18.2 (6.9) | 18.8 (7.9) | 0.94 | 83 |

Data presented as n (%) unless otherwise specified

**Supplementary table 7.** ICU support and therapeutic interventions at days 0 of ICU admission after IPTW and matching

|  | **Steroids** | **IL6-R antagonists** | **Both** | **P** | **SMD** |
| --- | --- | --- | --- | --- | --- |
| n¶ | 804.1 | 582.4 | 689.6 |  |  |
| **Code status change** |  |  |  | 0.62 | 0.137 |
| N/A | 17 ( 2.4) | 23 (4.0) | 11 (1.7) |  |  |
| No | 690 ( 94.9) | 531 (94.7) | 624 (94.8) |  |  |
| Yes | 20 ( 2.8) | 7 ( 1.3) | 23 ( 3.5) |  |  |
| **Code change** |  |  |  | 0.14 | 1.284 |
| Changed to 'no chest compression' (no CPR) -DNR | 12 ( 58.6) | 3 ( 40.1) | 0 ( 0) |  |  |
| Changed to both- DNI-DNR | 3 ( 12.8) | 0 ( 0) | 4 ( 18.0) |  |  |
| Changed to full support (full code) | 6 ( 28.6) | 4 ( 59.9) | 19 ( 82.0) |  |  |
| **Neuromuscular blocker** |  |  |  | 0.08 | 0.423 |
| Atracurium | 10 (1.6) | 6 (1.6) | 30 (6.4) |  |  |
| Cisatracurium | 75 (12.4) | 24 (6.2) | 26 (5.4) |  |  |
| Pancuronium | 4 ( 0.6) | 0 (0) | 0 (0) |  |  |
| Rocuronium | 117 ( 19.3) | 125 (32.6) | 125 (26.3) |  |  |
| Succinylcholine | 4 ( 0.6) | 5 ( 1.2) | 12 (2.5) |  |  |
| Vecuronium | 16 ( 2.7) | 12 ( 3.1) | 0 (0) |  |  |
| None | 370 ( 60.8) | 207 (54.0) | 277 (58.5) |  |  |
| **Stress ulcer prophylaxis** | 609 ( 77.6) | 367 (66.9) | 462 (70.2) | 0.14 | 0.159 |
| Esomeprazole | 22 ( 3.0) | 0 (0) | 0 (0) |  |  |
| Famotidine | 141 (19.4) | 110 (19.6) | 122 (18.7) |  |  |
| Lansoprazole | 20 ( 2.7) | 25 ( 4.5) | 39 (5.9) |  |  |
| Omeprazole | 92 (12.6) | 63 ( 11.2) | 98 (14.9) |  |  |
| Pantoprazole | 163 (22.4) | 58 (10.3) | 175 (26.7) |  |  |
| Ranitidine | 46 ( 6.3) | 0 ( 0.0) | 4 (0.6) |  |  |
| None | 210 (28.9) | 223 ( 39.8) | 176 (26.9) |  |  |
| Not Indicated | 30 ( 4.1) | 78 ( 14.0) | 41 (6.2) |  |  |
| Stress ulcer prophylaxis route (IV) | 325 (66.7) | 162 ( 62.4) | 287 (65.7) | 0.86 | 0.06 |
| Anticoagulant | 687 ( 85.4) | 494 ( 84.7) | 549 (79.6) | 0.36 | 0.102 |

Data presented as n (%) unless otherwise specified. N (%) may differ due to approximation
¶ Each individual gets its own weight that is used for further analysis. Multiplication by these weights can usually results in decimals
**Abbreviation:** CPR: Cardiopulmonary resuscitation; DNR: Do-not-resuscitate order; DNI: Do Not Intubate; IV: intravenous

**Supplementary table 8.** Distribution of Anakinra and Remdesivir doses

| Anakinra doses | N (%) |
| --- | --- |
| 0 | 851 (99) |
| 1 | 1 (0.1) |
| 2 | 3 (0.4) |
| 3 | 2 (0.2) |
| 4 | 2 (0.2) |
| 5 | 1 (0.1) |
| Remdesivir doses |  |
| 0 | 737 (85.7) |
| 1 | 24 (2.8) |
| 2 | 22 (2.6) |
| 3 | 21 (2.4) |
| 4 | 27 (3.1) |
| 5 | 14 (1.6) |
| 6 | 2 (0.2) |
| 7 | 2 (0.2) |
| 8 | 3 (0.4) |
| 9 | 1 (0.1) |
| 10 | 6 (0.7) |
| 11 | 1 (0.1) |

Data presented as n (%) unless otherwise specified

**Supplementary table 9.** When was the patient admitted to ICU with respect to hospital admission day.

|  | N (% of patients admitted to ICU) |  |  |
| --- | --- | --- | --- |
| Hospital day | **Steroid (n=589)** | **IL6-R antagonists (n=170)** | **Both (n=101)** |
| Day 0 | 437 (74.2)<sup>a</sup> | 96 (56.5)<sup>a</sup> | 59 (58.4)<sup>a</sup> |
| Day 1 | 71 (12.1) | 49 (28.8) | 31 (30.7) |
| Day 2 | 21 (3.57) | 9 (5.29) | 2 (1.98) |
| Day 3 | 27 (4.58) | 7 (4.12) | 5 (4.95) |
| Day 4 | 12 (2.04) | 1 (0.60) | 3 (2.97) |
| Day 5 | 2 (0.34) | 4 (2.35) | 0 (0.00) |
| Day 6 | 1 (0.17) | 1 (0.60) | 0 (0.00) |
| Day 7 | 13 (2.21) | 1 (0.60) | 2 (1.98) |
| Day 8 | 3 (0.51) | 0 (0.00) | 1 (0.99) |

<sup>a</sup> For many patients, the progression to critical illness occurs in a short period of time and this percentage also included patients who transferred from other hospitals

**Supplementary Table 10.**
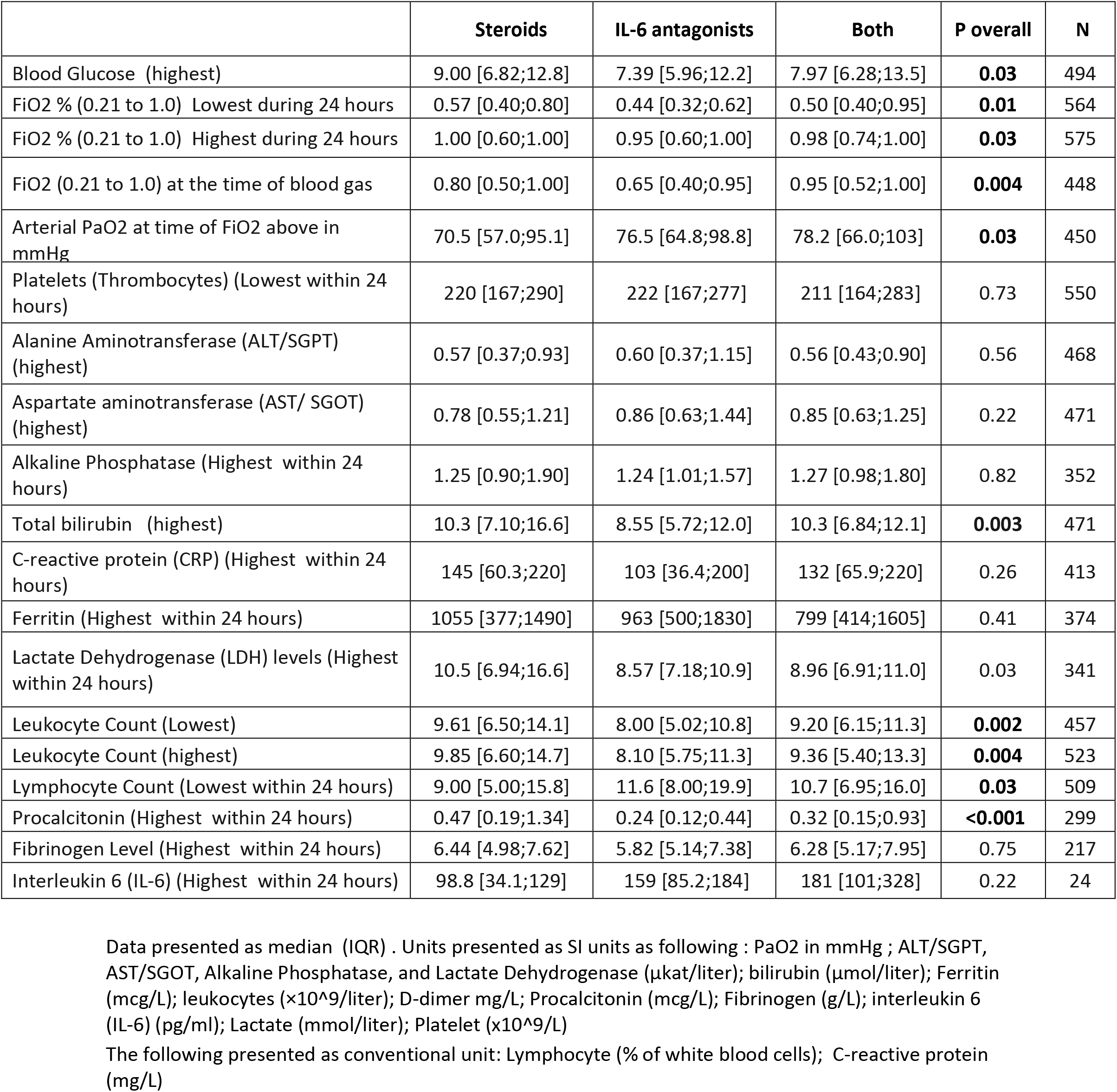
Laboratory parameters at ICU day 0 [unadjusted]

|  | **Steroids** | **IL-6 antagonists** | **Both** | **P overall** | **N** |
| --- | --- | --- | --- | --- | --- |
| Blood Glucose (highest) | 9.00 [6.82;12.8] | 7.39 [5.96;12.2] | 7.97 [6.28;13.5] | **0.03** | 494 |
| FiO2 % (0.21 to 1.0) Lowest during 24 hours | 0.57 [0.40;0.80] | 0.44 [0.32;0.62] | 0.50 [0.40;0.95] | **0.01** | 564 |
| FiO2 % (0.21 to 1.0) Highest during 24 hours | 1.00 [0.60;1.00] | 0.95 [0.60;1.00] | 0.98 [0.74;1.00] | **0.03** | 575 |
| FiO2 (0.21 to 1.0) at the time of blood gas | 0.80 [0.50;1.00] | 0.65 [0.40;0.95] | 0.95 [0.52;1.00] | **0.004** | 448 |
| Arterial PaO2 at time of FiO2 above in mmHg | 70.5 [57.0;95.1] | 76.5 [64.8;98.8] | 78.2 [66.0;103] | **0.03** | 450 |
| Platelets (Thrombocytes) (Lowest within 24 hours) | 220 [167;290] | 222 [167;277] | 211 [164;283] | 0.73 | 550 |
| Alanine Aminotransferase (ALT/SGPT) (highest) | 0.57 [0.37;0.93] | 0.60 [0.37;1.15] | 0.56 [0.43;0.90] | 0.56 | 468 |
| Aspartate aminotransferase (AST/ SGOT) (highest) | 0.78 [0.55;1.21] | 0.86 [0.63;1.44] | 0.85 [0.63;1.25] | 0.22 | 471 |
| Alkaline Phosphatase (Highest within 24 hours) | 1.25 [0.90;1.90] | 1.24 [1.01;1.57] | 1.27 [0.98;1.80] | 0.82 | 352 |
| Total bilirubin (highest) | 10.3 [7.10;16.6] | 8.55 [5.72;12.0] | 10.3 [6.84;12.1] | **0.003** | 471 |
| C-reactive protein (CRP) (Highest within 24 hours) | 145 [60.3;220] | 103 [36.4;200] | 132 [65.9;220] | 0.26 | 413 |
| Ferritin (Highest within 24 hours) | 1055 [377;1490] | 963 [500;1830] | 799 [414;1605] | 0.41 | 374 |
| Lactate Dehydrogenase (LDH) levels (Highest within 24 hours) | 10.5 [6.94;16.6] | 8.57 [7.18;10.9] | 8.96 [6.91;11.0] | 0.03 | 341 |
| Leukocyte Count (Lowest) | 9.61 [6.50;14.1] | 8.00 [5.02;10.8] | 9.20 [6.15;11.3] | **0.002** | 457 |
| Leukocyte Count (highest) | 9.85 [6.60;14.7] | 8.10 [5.75;11.3] | 9.36 [5.40;13.3] | **0.004** | 523 |
| Lymphocyte Count (Lowest within 24 hours) | 9.00 [5.00;15.8] | 11.6 [8.00;19.9] | 10.7 [6.95;16.0] | **0.03** | 509 |
| Procalcitonin (Highest within 24 hours) | 0.47 [0.19;1.34] | 0.24 [0.12;0.44] | 0.32 [0.15;0.93] | **<0.001** | 299 |
| Fibrinogen Level (Highest within 24 hours) | 6.44 [4.98;7.62] | 5.82 [5.14;7.38] | 6.28 [5.17;7.95] | 0.75 | 217 |
| Interleukin 6 (IL-6) (Highest within 24 hours) | 98.8 [34.1;129] | 159 [85.2;184] | 181 [101;328] | 0.22 | 24 |

Data presented as median (IQR) . Units presented as SI units as following : PaO2 in mmHg ; ALT/SGPT, AST/SGOT, Alkaline Phosphatase, and Lactate Dehydrogenase (μkat/liter); bilirubin (μmol/liter); Ferritin (mcg/L); leukocytes (×10<sup>9</sup>/liter); D-dimer mg/L; Procalcitonin (mcg/L); Fibrinogen (g/L); interleukin 6 (IL-6) (pg/ml); Lactate (mmol/liter); Platelet (x10<sup>9</sup>/L)
The following presented as conventional unit: Lymphocyte (% of white blood cells); C-reactive protein (mg/L)

**Supplementary Table 11.**
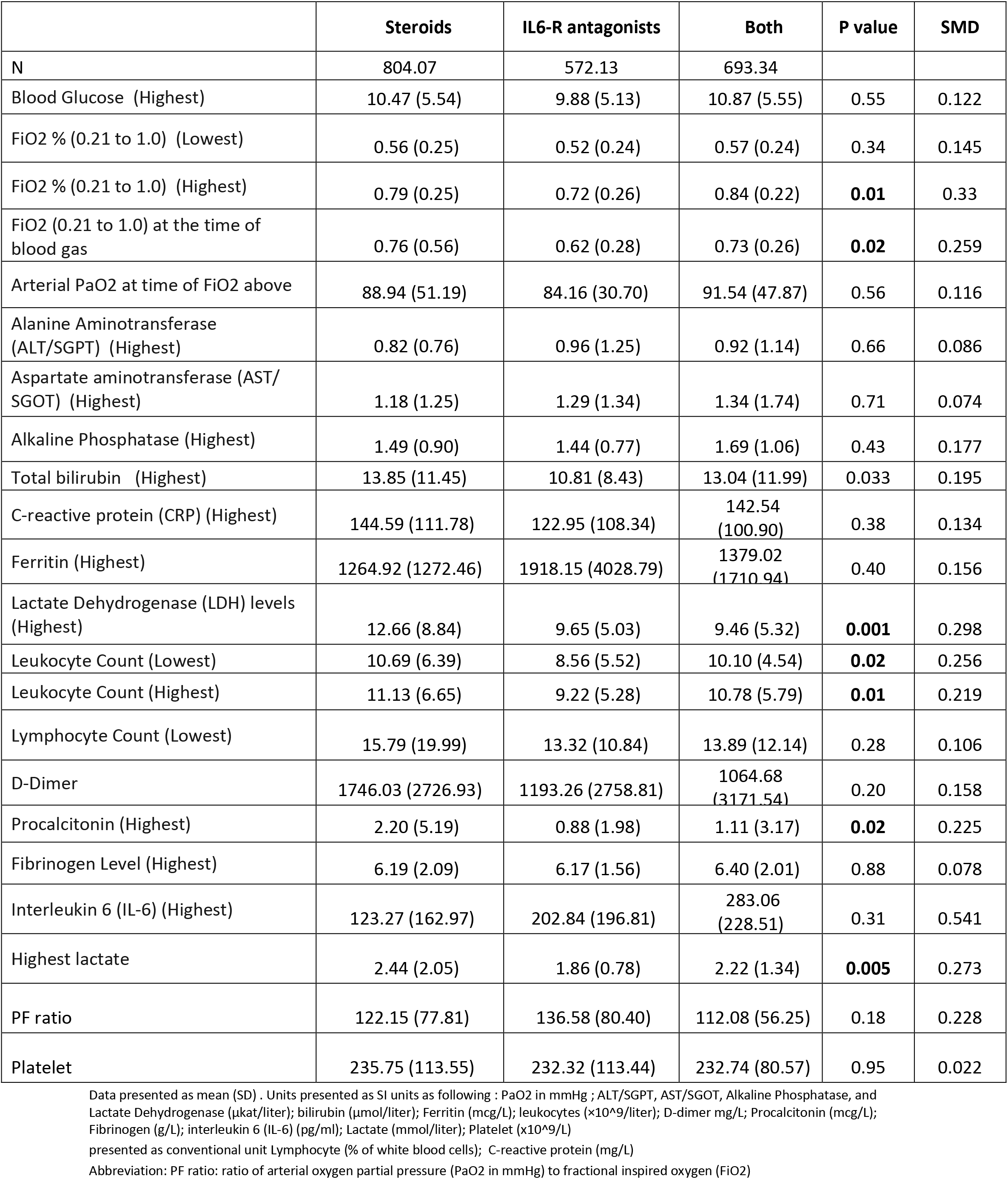
Laboratory parameters at ICU day 0 after adjustment of covariates

|  | **Steroids** | **IL6-R antagonists** | **Both** | **P value** | **SMD** |
| --- | --- | --- | --- | --- | --- |
| N | 804.07 | 572.13 | 693.34 |  |  |
| Blood Glucose (Highest) | 10.47 (5.54) | 9.88 (5.13) | 10.87 (5.55) | 0.55 | 0.122 |
| FiO2 % (0.21 to 1.0) (Lowest) | 0.56 (0.25) | 0.52 (0.24) | 0.57 (0.24) | 0.34 | 0.145 |
| FiO2 % (0.21 to 1.0) (Highest) | 0.79 (0.25) | 0.72 (0.26) | 0.84 (0.22) | **0.01** | 0.33 |
| FiO2 (0.21 to 1.0) at the time of blood gas | 0.76 (0.56) | 0.62 (0.28) | 0.73 (0.26) | **0.02** | 0.259 |
| Arterial PaO2 at time of FiO2 above | 88.94 (51.19) | 84.16 (30.70) | 91.54 (47.87) | 0.56 | 0.116 |
| Alanine Aminotransferase (ALT/SGPT) (Highest) | 0.82 (0.76) | 0.96 (1.25) | 0.92 (1.14) | 0.66 | 0.086 |
| Aspartate aminotransferase (AST/SGOT) (Highest) | 1.18 (1.25) | 1.29 (1.34) | 1.34 (1.74) | 0.71 | 0.074 |
| Alkaline Phosphatase (Highest) | 1.49 (0.90) | 1.44 (0.77) | 1.69 (1.06) | 0.43 | 0.177 |
| Total bilirubin (Highest) | 13.85 (11.45) | 10.81 (8.43) | 13.04 (11.99) | 0.033 | 0.195 |
| C-reactive protein (CRP) (Highest) | 144.59 (111.78) | 122.95 (108.34) | 142.54 (100.90) | 0.38 | 0.134 |
| Ferritin (Highest) | 1264.92 (1272.46) | 1918.15 (4028.79) | 1379.02 (1710.94) | 0.40 | 0.156 |
| Lactate Dehydrogenase (LDH) levels (Highest) | 12.66 (8.84) | 9.65 (5.03) | 9.46 (5.32) | **0.001** | 0.298 |
| Leukocyte Count (Lowest) | 10.69 (6.39) | 8.56 (5.52) | 10.10 (4.54) | **0.02** | 0.256 |
| Leukocyte Count (Highest) | 11.13 (6.65) | 9.22 (5.28) | 10.78 (5.79) | **0.01** | 0.219 |
| Lymphocyte Count (Lowest) | 15.79 (19.99) | 13.32 (10.84) | 13.89 (12.14) | 0.28 | 0.106 |
| D-Dimer | 1746.03 (2726.93) | 1193.26 (2758.81) | 1064.68 (3171.54) | 0.20 | 0.158 |
| Procalcitonin (Highest) | 2.20 (5.19) | 0.88 (1.98) | 1.11 (3.17) | **0.02** | 0.225 |
| Fibrinogen Level (Highest) | 6.19 (2.09) | 6.17 (1.56) | 6.40 (2.01) | 0.88 | 0.078 |
| Interleukin 6 (IL-6) (Highest) | 123.27 (162.97) | 202.84 (196.81) | 283.06 (228.51) | 0.31 | 0.541 |
| Highest lactate | 2.44 (2.05) | 1.86 (0.78) | 2.22 (1.34) | **0.005** | 0.273 |
| PF ratio | 122.15 (77.81) | 136.58 (80.40) | 112.08 (56.25) | 0.18 | 0.228 |
| Platelet | 235.75 (113.55) | 232.32 (113.44) | 232.74 (80.57) | 0.95 | 0.022 |

Data presented as mean (SD) . Units presented as SI units as following : PaO2 in mmHg ; ALT/SGPT, AST/SGOT, Alkaline Phosphatase, and Lactate Dehydrogenase (μkat/liter); bilirubin (μmol/liter); Ferritin (mcg/L); leukocytes (×10<sup>9</sup>/liter); D-dimer mg/L; Procalcitonin (mcg/L); Fibrinogen (g/L); interleukin 6 (IL-6) (pg/ml); Lactate (mmol/liter); Platelet (×10<sup>9</sup>/L)
presented as conventional unit Lymphocyte (% of white blood cells); C-reactive protein (mg/L)
Abbreviation: PF ratio: ratio of arterial oxygen partial pressure (PaO2 in mmHg) to fractional inspired oxygen (FiO2)

**Supplementary Table 12:** Clinical outcomes before matching

|  | **Steroids** | **IL6-R antagonists** | **Both** | **P overall** |
| --- | --- | --- | --- | --- |
|  | **N=589** | **N=170** | **N=101** |  |
| **ICU discharge status** |  |  |  | <0.001 |
| Alive | 296 (50.3) | 118 (69.4) | 51 (50.4) |  |
| Deceased | 287 (48.7) | 50 (30.6) | 53 (49.5) |  |
| **Hospital discharge status** |  |  |  | <0.001 |
| Alive | 294 (49.9) | 119 (70) | 47 (46.5) |  |
| Deceased | 295 (50.1) | 51 (30) | 54 (53.5) |  |
| **Ventilation free days, median IQR** | 18 [10;22] | 16.2 [7.50;21.4] | 14 [6.4;21] | 0.03 |
| **Duration of mechanical ventilation (Survivors only)** | 10.42 [6; 20.5] | 13.75 [7.8; 22.1] | 14.2 [9.6; 20.8] | 0.16 |
| **Total ICU length of Stay, median IQR** | 13.8 [8.00;22.0] | 13.4 [8.14;23.8] | 17.0 [9.38;24.3] | 0.08 |
| **Total ICU length of Stay in survivor, median IQR** | 15.2 [9.00;25.0] | 14.0 [8.89;24.0] | 18.0 [13.1;25.6] | 0.22 |
| **Total hospital length of Stay, median IQR** | 18 [10.6;28.4] | 19 [11.0;31] | 23.9 [13.1;33] | 0.05 |
| Statistical analysis was performed using Chi-square test of independent for ICU discharge and Kruskal-Wallis test for continuous variables<br>Data presented as n (%) unless otherwise specified |  |  |  |  |

**Supplementary table 13:** Association between medication use and mortality after adjustment stratified by steroid dose

| *Predictors* | *Adj Odds Ratio* | *95% CI* | *p* |
| --- | --- | --- | --- |
| **IL-6R antagonist** | *Reference* |  |  |
| **Steroid Low-dose** | 1.28 | 0.66 – 2.51 | 0.46 |
| **Steroid High-dose** | 1.73 | 0.75 – 4.04 | 0.20 |
| Age (1 year increase) | 1.05 | 1.03 – 1.07 | **<0.001** |
| Weight (1 Kg increase) | 1.00 | 0.99 – 1.01 | 0.90 |
| Lowest FiO2 | 3.20 | 1.26 – 8.27 | **0.02** |
| **Gender** |  |  |  |
| Female | *Reference* |  |  |
| Male | 1.32 | 0.82 – 2.15 | 0.26 |
| **Neuromuscular blocker** |  |  |  |
| Yes | *Reference* |  |  |
| No | 0.87 | 0.40 – 1.92 | 0.73 |
| **Vasopressors** |  |  |  |
| Yes | *Reference* |  |  |
| No | 0.88 | 0.42 – 1.82 | 0.72 |
| **Therapeutic anticoagulation** |  |  |  |
| No | *Reference* |  |  |
| Yes | 1.41 | 0.80 – 2.46 | 0.23 |
| **Any anticoagulation** |  |  |  |
| Yes | *Reference* |  |  |
| No | 0.60 | 0.31 – 1.16 | 0.13 |
| **Ethnicity** |  |  |  |
| Hispanic | *Reference* |  |  |
| Non-Hispanic | 1.41 | 0.79 – 2.55 | 0.25 |
| **Hydroxychloroquine** |  |  |  |
| Yes | *Reference* |  |  |
| No | 1.16 | 0.62 – 2.16 | 0.64 |
| **Antivirals** |  |  |  |
| Yes | *Reference* |  |  |
| No | 0.82 | 0.44 – 1.54 | 0.53 |
| **Azithromycin** |  |  |  |
| Yes | *Reference* |  |  |
| No | 1.32 | 0.78 – 2.22 | 0.30 |
| **ARDS grade** |  |  |  |
| No | *Reference* |  |  |
| Mild (P:F 200-300) | 0.42 | 0.07 – 2.20 | 0.32 |
| Moderate (P:F 100-199) | 0.61 | 0.25 – 1.46 | 0.27 |
| Severe (P:F< 100) | 0.98 | 0.48 – 1.97 | 0.95 |
| **Cardiovascular diseases** |  |  |  |
| No | *Reference* |  |  |
| Yes | 2.27 | 1.29 – 4.08 | **0.01** |
| **Asthma/COPD** |  |  |  |
| No | *Reference* |  |  |
| Yes | 0.94 | 0.51 – 1.72 | 0.84 |

The model was adjusted for the same variables used for IPTW.
Abbreviation: OR: odds ratio; CI: confidence interval; IPTW: Inverse probability

**Supplementary Figure 1.**
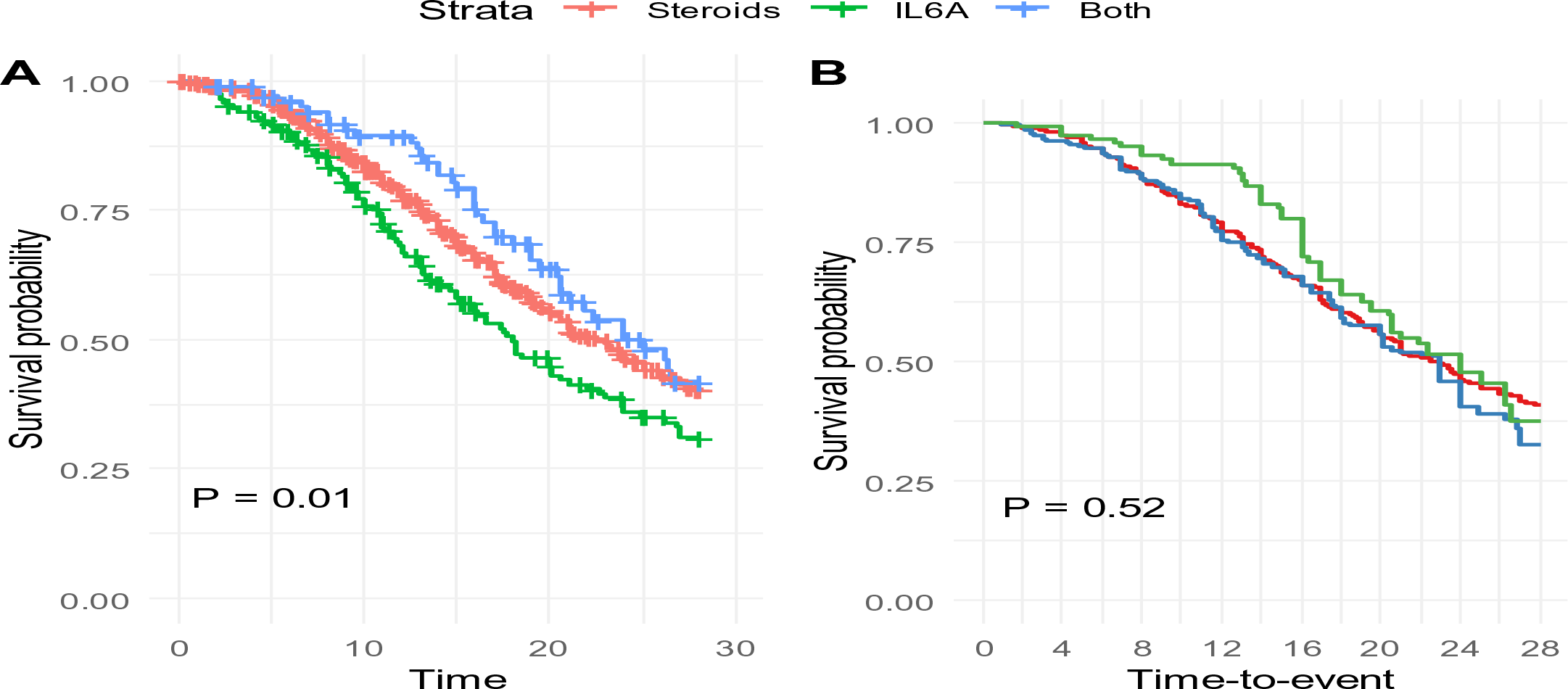
Unweighted and weighted Kaplan–Maier estimates for association between treatment ICU LOS

**Supplementary Figure 2.**
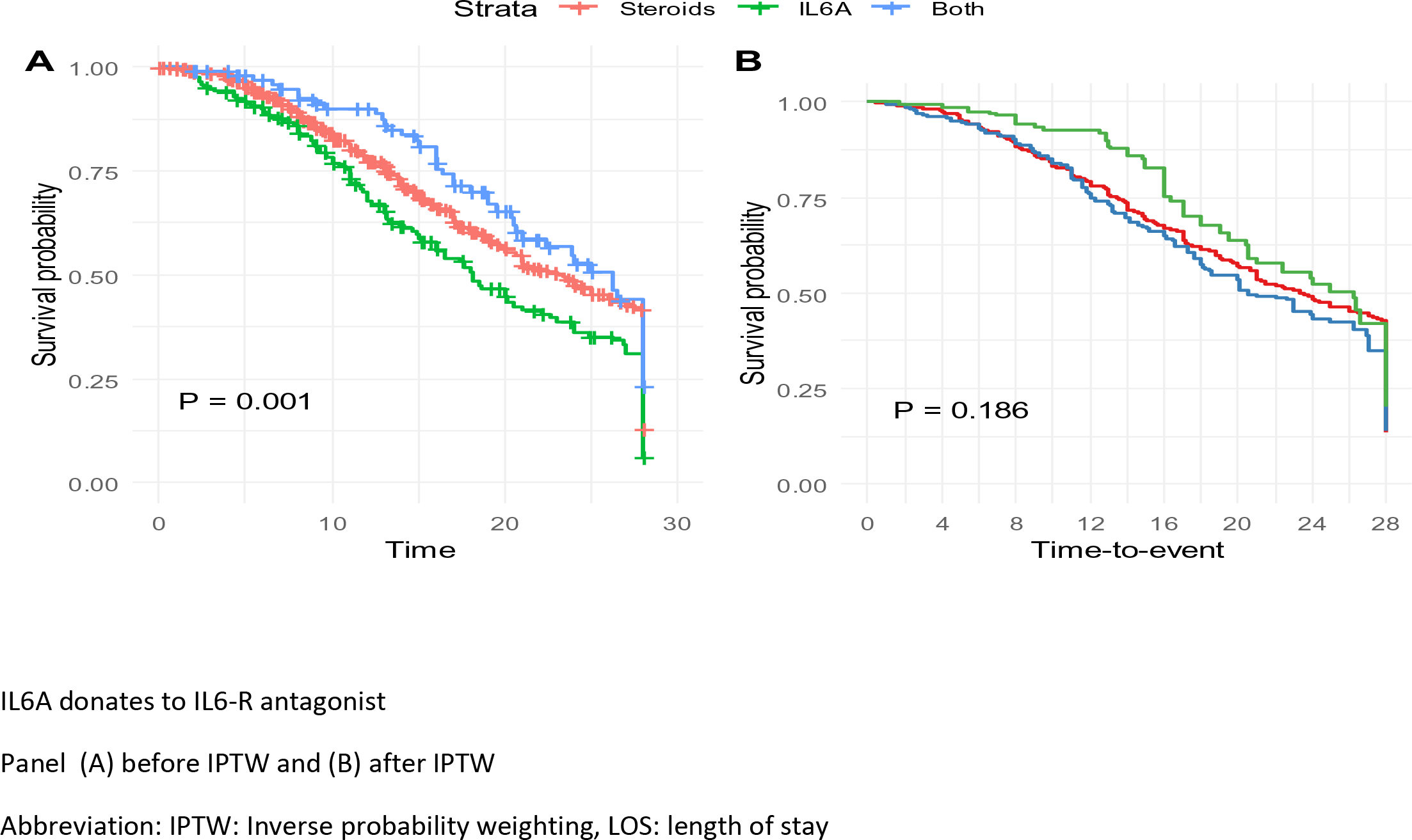
Unweighted and weighted Kaplan–Maier estimates for association between treatment and hospital LOS

**Supplementary table 14.**
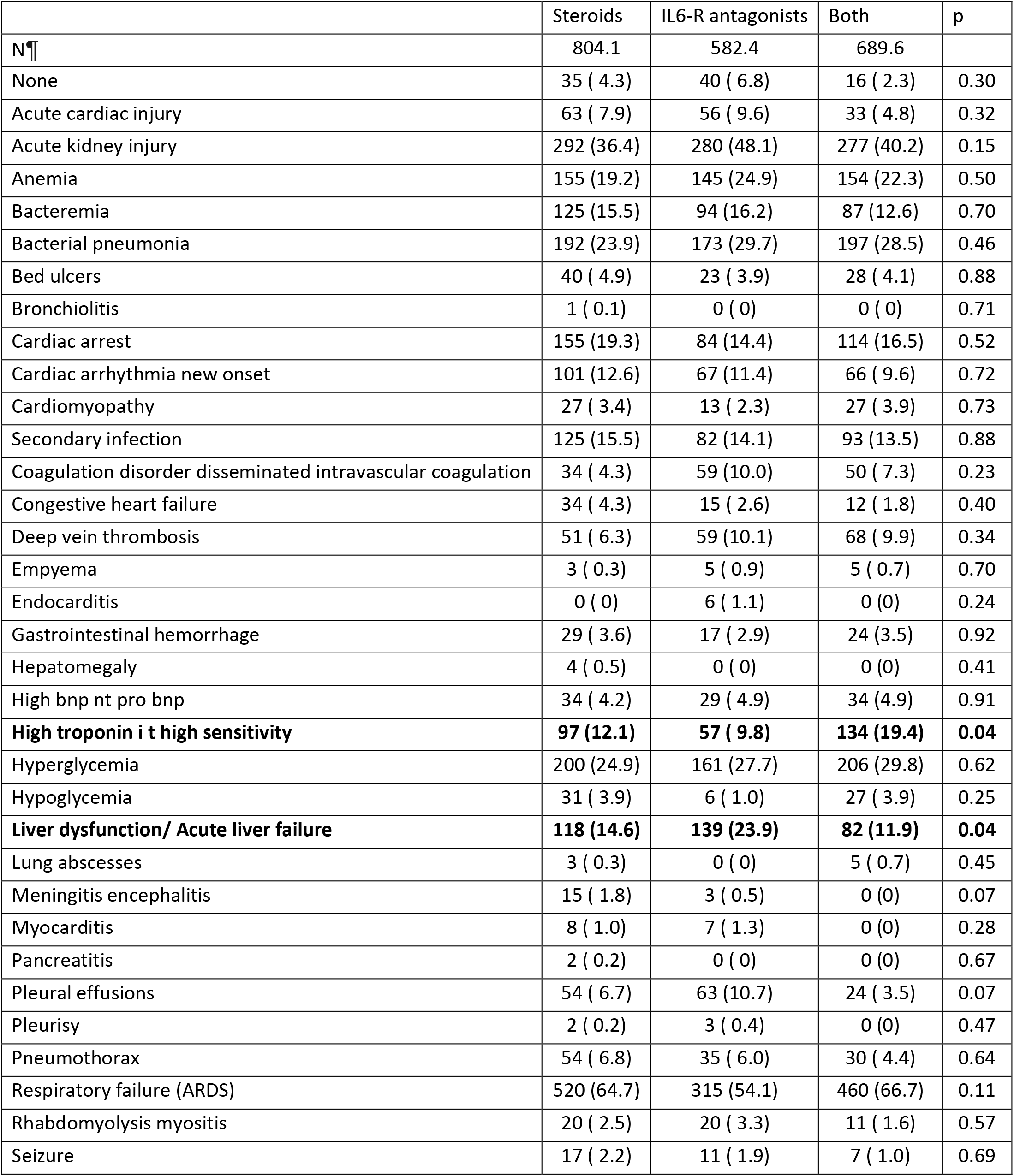
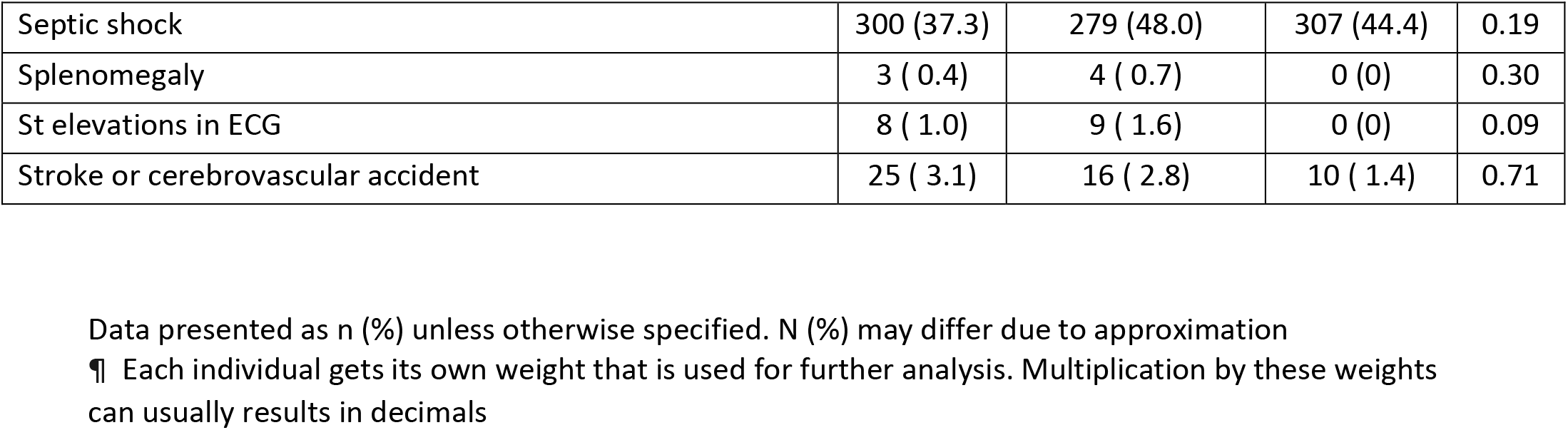
Documented complication during hospitalization after adjusting for baseline covariates

|  | Steroids | IL6-R antagonists | Both | p |
| --- | --- | --- | --- | --- |
| N | 804.1 | 582.4 | 689.6 |  |
| None | 35 ( 4.3) | 40 ( 6.8) | 16 ( 2.3) | 0.30 |
| Acute cardiac injury | 63 ( 7.9) | 56 ( 9.6) | 33 ( 4.8) | 0.32 |
| Acute kidney injury | 292 (36.4) | 280 (48.1) | 277 (40.2) | 0.15 |
| Anemia | 155 (19.2) | 145 (24.9) | 154 (22.3) | 0.50 |
| Bacteremia | 125 (15.5) | 94 (16.2) | 87 (12.6) | 0.70 |
| Bacterial pneumonia | 192 (23.9) | 173 (29.7) | 197 (28.5) | 0.46 |
| Bed ulcers | 40 ( 4.9) | 23 ( 3.9) | 28 ( 4.1) | 0.88 |
| Bronchiolitis | 1 ( 0.1) | 0 ( 0) | 0 ( 0) | 0.71 |
| Cardiac arrest | 155 (19.3) | 84 (14.4) | 114 (16.5) | 0.52 |
| Cardiac arrhythmia new onset | 101 (12.6) | 67 (11.4) | 66 ( 9.6) | 0.72 |
| Cardiomyopathy | 27 ( 3.4) | 13 ( 2.3) | 27 ( 3.9) | 0.73 |
| Secondary infection | 125 (15.5) | 82 (14.1) | 93 (13.5) | 0.88 |
| Coagulation disorder disseminated intravascular coagulation | 34 ( 4.3) | 59 (10.0) | 50 ( 7.3) | 0.23 |
| Congestive heart failure | 34 ( 4.3) | 15 ( 2.6) | 12 ( 1.8) | 0.40 |
| Deep vein thrombosis | 51 ( 6.3) | 59 (10.1) | 68 ( 9.9) | 0.34 |
| Empyema | 3 ( 0.3) | 5 ( 0.9) | 5 ( 0.7) | 0.70 |
| Endocarditis | 0 ( 0) | 6 ( 1.1) | 0 (0) | 0.24 |
| Gastrointestinal hemorrhage | 29 ( 3.6) | 17 ( 2.9) | 24 (3.5) | 0.92 |
| Hepatomegaly | 4 ( 0.5) | 0 ( 0) | 0 (0) | 0.41 |
| High bnp nt pro bnp | 34 ( 4.2) | 29 ( 4.9) | 34 (4.9) | 0.91 |
| **High troponin i t high sensitivity** | **97 (12.1)** | **57 ( 9.8)** | **134 (19.4)** | **0.04** |
| Hyperglycemia | 200 (24.9) | 161 (27.7) | 206 (29.8) | 0.62 |
| Hypoglycemia | 31 ( 3.9) | 6 ( 1.0) | 27 ( 3.9) | 0.25 |
| **Liver dysfunction/ Acute liver failure** | **118 (14.6)** | **139 (23.9)** | **82 (11.9)** | **0.04** |
| Lung abscesses | 3 ( 0.3) | 0 ( 0) | 5 ( 0.7) | 0.45 |
| Meningitis encephalitis | 15 ( 1.8) | 3 ( 0.5) | 0 (0) | 0.07 |
| Myocarditis | 8 ( 1.0) | 7 ( 1.3) | 0 (0) | 0.28 |
| Pancreatitis | 2 ( 0.2) | 0 ( 0) | 0 (0) | 0.67 |
| Pleural effusions | 54 ( 6.7) | 63 (10.7) | 24 ( 3.5) | 0.07 |
| Pleurisy | 2 ( 0.2) | 3 ( 0.4) | 0 (0) | 0.47 |
| Pneumothorax | 54 ( 6.8) | 35 ( 6.0) | 30 ( 4.4) | 0.64 |
| Respiratory failure (ARDS) | 520 (64.7) | 315 (54.1) | 460 (66.7) | 0.11 |
| Rhabdomyolysis myositis | 20 ( 2.5) | 20 ( 3.3) | 11 ( 1.6) | 0.57 |
| Seizure | 17 ( 2.2) | 11 ( 1.9) | 7 ( 1.0) | 0.69 |
| Septic shock | 300 (37.3) | 279 (48.0) | 307 (44.4) | 0.19 |
| Splenomegaly | 3 ( 0.4) | 4 ( 0.7) | 0 (0) | 0.30 |
| St elevations in ECG | 8 ( 1.0) | 9 ( 1.6) | 0 (0) | 0.09 |
| Stroke or cerebrovascular accident | 25 ( 3.1) | 16 ( 2.8) | 10 ( 1.4) | 0.71 |

Data presented as n (%) unless otherwise specified. N (%) may differ due to approximation
¶ Each individual gets its own weight that is used for further analysis. Multiplication by these weights can usually results in decimals

**Supplementary table 15:** Causative microbiology and rate of secondary infections across the groups

|  | Steroids | IL-6 antagonists | Both | P |
| --- | --- | --- | --- | --- |
| Respiratory Viral Panel (RVP) PCR Positive¶ | 1 (0.2) | 2 (1.2) | 0 (0) | 0.72 |
| Hepatitis B Positive | 9 (1.5) | 9 (5.3) | 2 (1.9) | 0.02 |
| Hepatitis C Positive | 11 (1.9) | 1 (0) | 1 (0.9) | 0.62 |
| Cytomegalovirus PCR Positive | 2 (0.3) | 0 (0) | 1 (0.9) | 0.40 |
| Legionella Urine Ag Positive | 0 (0) | 1 (0.6) | 0 (0) | 0.32 |
| Blood Cultures Positive # | 93 (21.9) | 27 (17.2) | 17 (19.3) | 0.44 |
| Which Hospital Day(s) Positive Blood Cultures Performed? Median [IQR] | 7 [0.25;14] | 14[1;22] | 10 [0.75;13] | 0.14 |
| Sputum Culture Positive § | 167 (50.6) | 65 (53.3) | 34 (45.9) | 0.61 |
| Which Hospital Day(s) Positive Sputum Culture Performed? Median [IQR] | 4 [1;11] | 5[1.5;9] | 5 [1;13.5] | 0.87 |
| Urine Culture Positive? * | 74 (29.2) | 23 (20.2) | 11 (19.6) | 0.10 |
| Overall Bacterial super infection | 87 (14.8) | 27 (15.9) | 15 (14.9) | 0.90 |

¶ performed in 127 steroids group, 85 IL6-R antagonist group, 33 combination group

**Supplementary table 16.**
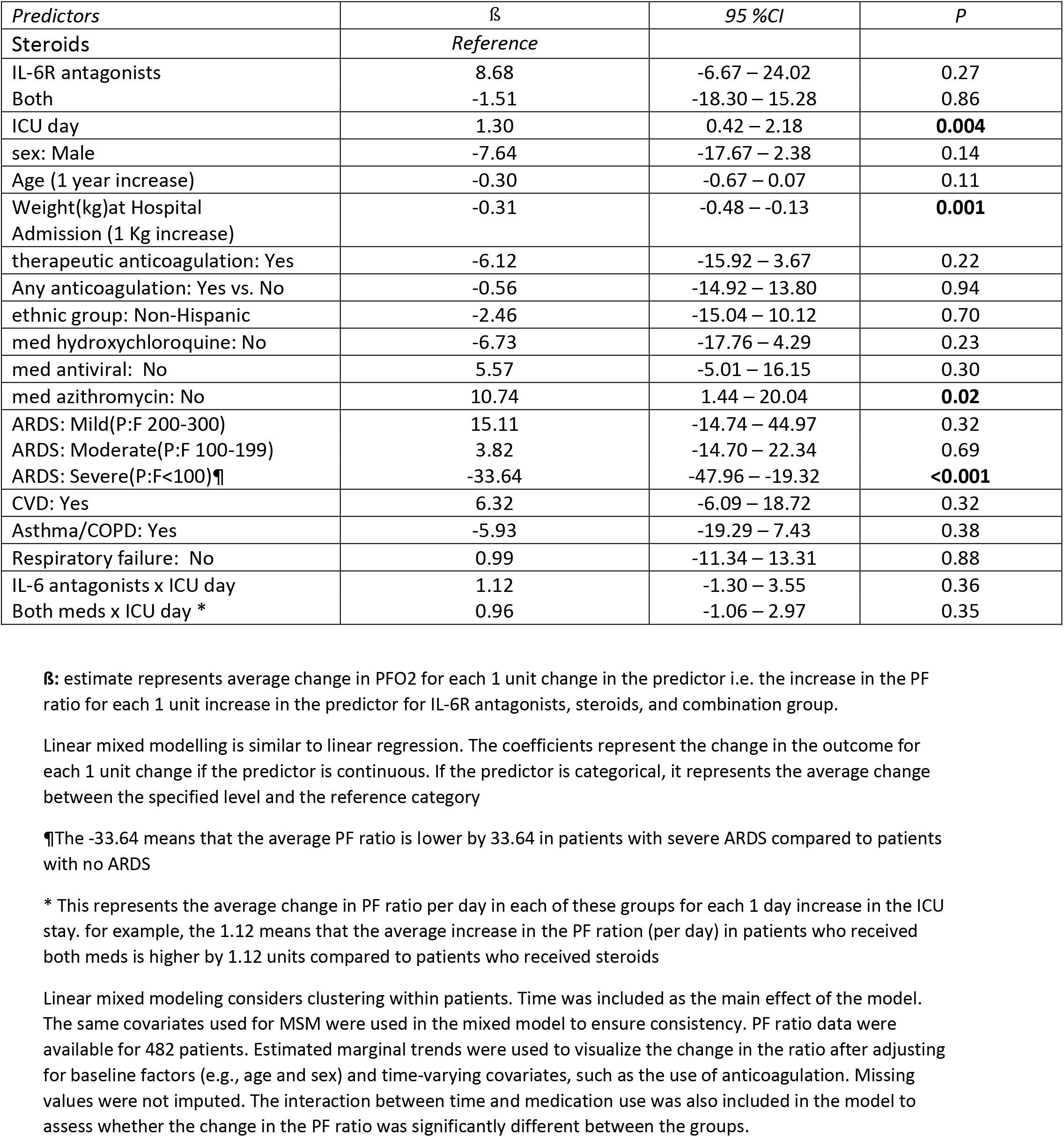
Linear mixed modelling of PF ratio. Linear mixed model and linear marginal trends were used to assess changes in the PF ratio after adjusting for the same covariates and showed that the use of IL-6RA or combination therapy was not associated with significantly different changes in the PF ratio compared with the use of steroids alone.

| Predictors | $\beta$ | 95 %CI | P |
| --- | --- | --- | --- |
| Steroids | Reference |  |  |
| IL-6R antagonists | 8.68 | -6.67 – 24.02 | 0.27 |
| Both | -1.51 | -18.30 – 15.28 | 0.86 |
| ICU day | 1.30 | 0.42 – 2.18 | **0.004** |
| sex: Male | -7.64 | -17.67 – 2.38 | 0.14 |
| Age (1 year increase) | -0.30 | -0.67 – 0.07 | 0.11 |
| Weight(kg)at Hospital Admission (1 Kg increase) | -0.31 | -0.48 – -0.13 | **0.001** |
| therapeutic anticoagulation: Yes | -6.12 | -15.92 – 3.67 | 0.22 |
| Any anticoagulation: Yes vs. No | -0.56 | -14.92 – 13.80 | 0.94 |
| ethnic group: Non-Hispanic | -2.46 | -15.04 – 10.12 | 0.70 |
| med hydroxychloroquine: No | -6.73 | -17.76 – 4.29 | 0.23 |
| med antiviral: No | 5.57 | -5.01 – 16.15 | 0.30 |
| med azithromycin: No | 10.74 | 1.44 – 20.04 | **0.02** |
| ARDS: Mild(P:F 200-300) | 15.11 | -14.74 – 44.97 | 0.32 |
| ARDS: Moderate(P:F 100-199) | 3.82 | -14.70 – 22.34 | 0.69 |
| ARDS: Severe(P:F<100)¶ | -33.64 | -47.96 – -19.32 | **<0.001** |
| CVD: Yes | 6.32 | -6.09 – 18.72 | 0.32 |
| Asthma/COPD: Yes | -5.93 | -19.29 – 7.43 | 0.38 |
| Respiratory failure: No | 0.99 | -11.34 – 13.31 | 0.88 |
| IL-6 antagonists x ICU day | 1.12 | -1.30 – 3.55 | 0.36 |
| Both meds x ICU day * | 0.96 | -1.06 – 2.97 | 0.35 |

$\beta$ : estimate represents average change in PFO2 for each 1 unit change in the predictor i.e. the increase in the PF ratio for each 1 unit increase in the predictor for IL-6R antagonists, steroids, and combination group.
Linear mixed modelling is similar to linear regression. The coefficients represent the change in the outcome for each 1 unit change if the predictor is continuous. If the predictor is categorical, it represents the average change between the specified level and the reference category
¶The -33.64 means that the average PF ratio is lower by 33.64 in patients with severe ARDS compared to patients with no ARDS
\* This represents the average change in PF ratio per day in each of these groups for each 1 day increase in the ICU stay. for example, the 1.12 means that the average increase in the PF ration (per day) in patients who received both meds is higher by 1.12 units compared to patients who received steroids
Linear mixed modeling considers clustering within patients. Time was included as the main effect of the model. The same covariates used for MSM were used in the mixed model to ensure consistency. PF ratio data were available for 482 patients. Estimated marginal trends were used to visualize the change in the ratio after adjusting for baseline factors (e.g., age and sex) and time-varying covariates, such as the use of anticoagulation. Missing values were not imputed. The interaction between time and medication use was also included in the model to assess whether the change in the PF ratio was significantly different between the groups.

**Supplementary Figure 3.**
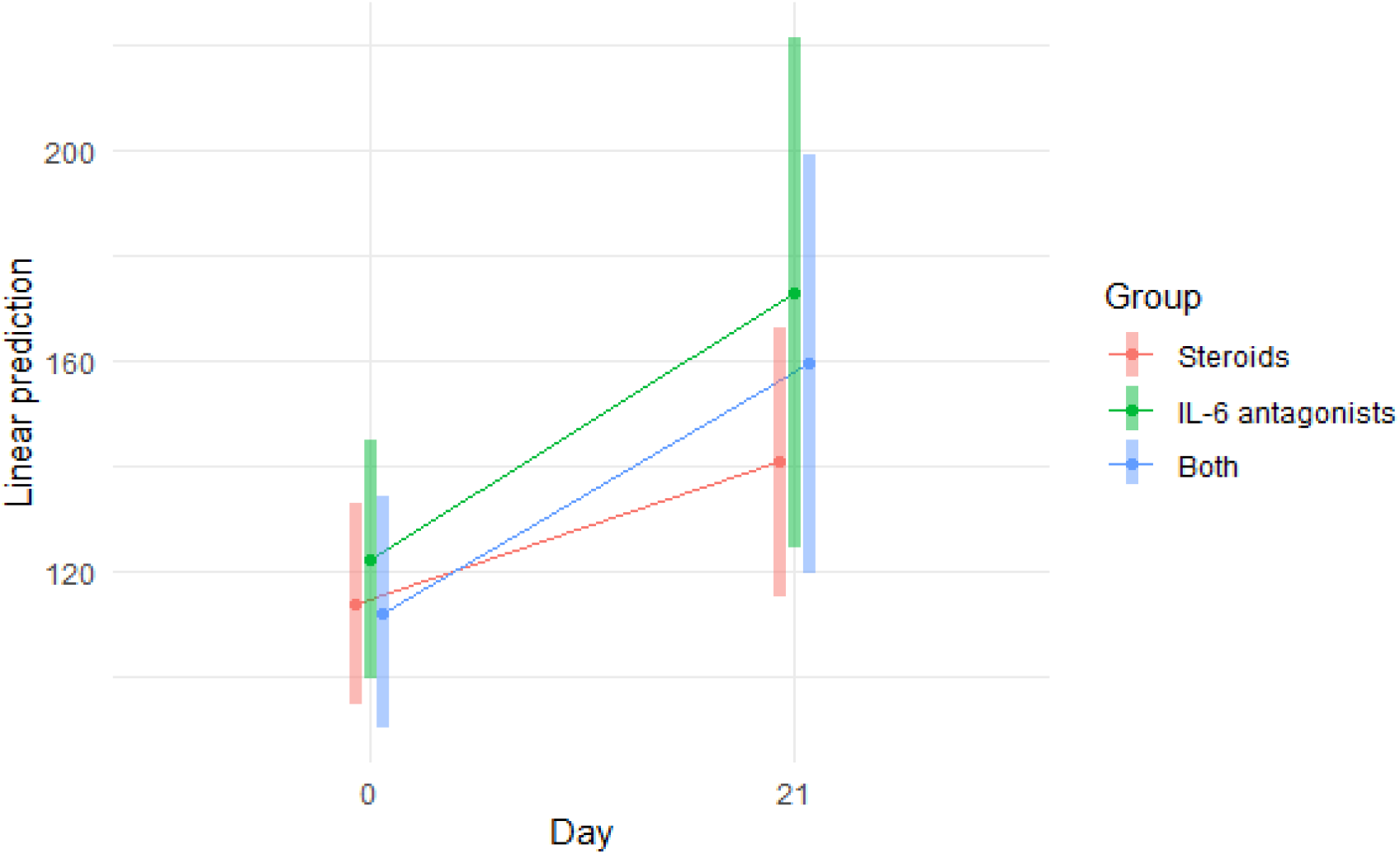
Estimated marginal trend for the change in PF ratio across groups.

**Supplementary table 17:** Change in lab values. Linear mixed modelling was used to assess whether the change in lab values across time was significantly different between groups. The interaction between time and medication was included in the model. The same variables used for IPTW were used for the analysis. Only results for the interaction are shown.

| | $\beta$ | SE | t | p |
| --- | --- | --- | --- | --- |
| **CRP (n = 247)** |  |  |  |  |
| IL-6R antagonist | -0.98 | 1.8 | -0.55 | 0.58 |
| Both | -3.39 | 1.64 | -2.07 | **0.04** |
| **Lowest lymphocyte (n = 372)** |  |  |  |  |
| IL-6R antagonist | 0.19 | 0.25 | 0.76 | 0.45 |
| Both | 0.13 | 0.23 | 0.59 | 0.56 |
| **Lowest leukocyte (n = 352)** |  |  |  |  |
| IL-6R antagonist | 0.2 | 0.1 | 1.97 | **0.05** |
| Both | 0.07 | 0.09 | 0.78 | 0.43 |
| **Highest LDH (n = 205)** |  |  |  |  |
| IL-6R antagonist | 0.01 | 0.12 | 0.12 | 0.91 |
| Both | 0.02 | 0.12 | 0.19 | 0.85 |
| **Ferritin (n = 220)** |  |  |  |  |
| IL-6R antagonist | -51.65 | 24.67 | -2.09 | **0.04** |
| Both | -32.19 | 19.77 | -1.63 | 0.10 |
| Data for IL-6 was not sufficient<br>N represents the number of available patients for the analysis |  |  |  |  |

## Supplementary File 2: Collaborative Co-Author List

¥ On behalf of the Society of Critical Care Medicine Discovery Viral Infection and Respiratory Illness Universal Study (VIRUS): COVID-19 Registry Investigator Group

**<u>Belgium</u>**

Centre Hospitalier Jolimont: Jean-Baptiste Mesland, Pierre Henin, Hélène Petre, Isabelle Buelens, Anne-Catherine Gerard

The Brugmann University hospital, Bruxelles: Philippe Clevenbergh

**<u>Bosnia and Herzegovina</u>**

University Clinical Hospital, Mostar, Bosnia and Herzegovina: Dragana Markotić, Ivana Bošnjak

**<u>Columbia</u>**

Clinica Medical SAS: Oscar Y Gavidia, Felipe Pachon, Yeimy A Sanchez

**<u>Croatia</u>**

Clinical Hospital Center Rijeka, Rijeka, Croatia: Danijel knežević

**<u>Egypt</u>**

Helwan University: Mohamed El Kassas, Mohamed Badr, Ahmed Tawheed, Ahmed Tawheed, Hend Yahia

**<u>Hondurus</u>**

Honduras Medical Center: Jose Luis Ramos Coello, Guillermo Perez, Ana Karen Vallecillo Lizardo, Gabina María Reyes Guillen, Helin Archaga Soto

**<u>Hungary</u>**

Uzsoki Teaching Hospital: Csaba Kopitkó, Ágnes Bencze, István Méhész, Zsófia Gerendai,

**<u>India</u>**

Jawaharlal Institute of Postgraduate Medical Education and Research, Pondicherry: Anusha Cherian, Sreejith Parameswaran, Magesh Parthiban, Menu Priya A.

Maulana Azad Medical College and Lok Nayak hospital: Mradul Kumar Daga, Munisha Agarwal, Ishan Rohtagi

**<u>Japan</u>**

Center Hospital of the National Center for Global Health and Medicine: Wataru Matsuda, Reina Suzuki

Sapporo City General Hospital: Yuki Itagaki, Akira Kodate, Reina Suzuki, Akira Kodate, Yuki Takahashi, Koyo Moriki

Tokyo Medical and Dental University: Hidenobu Shigemitsu, Yuka Mishima, Nobuyuki Nosaka, Michio Nagashima

Hiroshima University: Michihito Kyo

**<u>Mexico</u>**

Hospital Universitario, Universidad Autonoma de Nuevo León: Rene Rodriguez-Gutierrez, Jose Gerardo Gonzalez-Gonzalez, Alejandro Salcido-Montenegro, Adrian Camacho-Ortiz

**<u>Pakistan</u>**

Dow University Hospital: Muhammad Sohaib Asghar, Mashaal Syed, Syed Anosh Ali Naqvi

**<u>Russia</u>**

Kuban State Medical University with affiliation Territorial Hospital #2: Igor Borisovich Zabolotskikh, Konstantin Dmitrievich Zybin, Sergey Vasilevich Sinkov, Tatiana Sergeevna Musaeva

**<u>Saudi Arabia</u>**

King Saud University: Mohammed A Almazyad, Mohammed I Alarifi, Jara M Macarambon, Ahmad Abdullah Bukhari, Hussain A. Albahrani, Kazi N Asfina, Kaltham M Aldossary

King Saud bin Abdulaziz University for Health Sciences and King Abdullah International Medical Research Center: Yaseen M Arabi, Sheryl Ann Abdukahil

King Fahad Armed Forces Hospital: Razan K Alamoudi, Hassan M. AlSharif, Sarah A. Almazwaghi, Mohammed S Elsakran, Mohamed A Aid, Mouaz A Darwich, Omnia M Hagag, Salah A Ali, Alona rocacorba, Kathrine Supeña, Efren Ray Juane, Jenalyn Medina, Jowany Baduria

King Faisal Specialist Hospital & Research Centre-Riyadh: Marwa Ridha Amer, Mohammed Abdullah Bawazeer, Khalid Maghrabi, Abid Shahzad Butt, Talal I. Dahhan, Eiad Kseibi, Syed Moazzum Khurshid, Muath Rabee, Mohammed Abujazar, Razan Alghunaim, Maal Abualkhair, Abeer Turki AlFirm, Ali Al-Janoubi

**<u>Serbia</u>**

UMC Zvezdara, Belgrad: Bojan Kovacevic, Jovana Bojicic

Institute for Pulmonary Diseases of Vojvodina, Sremska Kamenica: Ana Andrijevic, Srdjan Gavrilovic, Vladimir Carapic

**<u>Spain</u>**

Hospital Universitario La Paz: Santiago Y. Teruel, Belen C. Martin, Santiago Y. Teruel

**<u>United States</u>**

Boston University School of Medicine, Boston, MA: Allan J. Walkey, Sushrut S. Waikar, Michael A. Garcia, Mia Colona, Zoe Kibbelaar, Michael Leong, Daniel Wallman, Kanupriya Soni, Jennifer Maccarone, Joshua Gilman, Ycar Devis, Joseph Chung, Munizay Paracha, David N. Lumelsky, Madeline DiLorenzo, Najla Abdurrahman, Shelsey Johnson

Albany Medical Center: Suzanne Barry, Christopher Woll, Gregory Wu, Erin Carrole, Kathryn Burke, Mustafa Mohammed

AnMed Health: Abhijit A Raval, Andrea Franks

Ascension St.Vincent Hospital: Anmol Kharbanda, Sunil Jhajhria, Zachary Fyffe

Ascension/St. Thomas Research Institute West Campus: Stephen Capizzi, Bethany Alicie, Martha

Green, Lori Crockarell, Amelia Drennan, Kathleen Dubuque, Tonya Fambrough, Nikole Gasaway, Briana Krantz, Peiman Nebi, Jan Orga, Margaret Serfass, Alina Simion, Kimberly Warren, Cassie Wheeler, CJ Woolman

Augusta Health: Andrew S. Moyer, George M. Verghese

Augusta University Medical Center: Andrea Sikora Newsome, Christy C. Forehand, Rebecca Bruning, Timothy W. Jones

Banner University Medical Center-Tucson: Jarrod M Mosier, Karen Lutrick, Beth Salvagio Campbell, Cathleen Wilson, Patrick Rivers, Jonathan Brinks, Mokenge Ndiva Mongoh, Boris Gilson

Baylor College of Medicine, Baylor St. Lukes Medical Center: Christopher M Howard, Cameron McBride, Jocelyn Abraham, Orlando Garner, Katherine Richards, Keegan Collins, Preethi Antony, Sindhu Mathew

Baylor Scott & White Health: Valerie C. Danesh, Gueorgui Dubrocq, Amber L. Davis, Marissa J Hammers, ill M. McGahey, Amanda C. Farris, Elisa Priest, Robyn Korsmo, Lorie Fares, Kathy Skiles, Susan M. Shor, Kenya Burns, Corrie A Dowell, Gabriela “Hope” Gonzales, Melody Flores, Lindsay Newman, Debora A Wilk, Jason Ettlinger, Jaccallene Bomar, Himani Darji, Alejandro Arroliga, Alejandro C Arroliga, Corrie A. Dowell, Gabriela Hope Conzales, Melody Flores, Lindsay Newman, Debora A. Wilk, Jason Ettlinger, Himani Darji, Jaccallene Bomar

Beth Israel Deaconess Medical Center: Valerie M. Banner-Goodspeed, Somnath Bose, Lauren E. Kelly, Melisa Joseph, Marie McGourty, Krystal Capers, Benjamin Hoenig, Maria C. Karamourtopoulos, Anica C. Law, Elias N. Baedorf Kassis

Cedars Sinai Medical Center: Pooja A. Nawathe, Isabel Pedraza, Jennifer Tsing, Karen Carr, Anila Chaudhary, Kathleen Guglielmino

Chambersburg Hospital: Raghavendra Tirupathi, Alymer Tang, Arshad Safi, Cindy Green, Jackie Newell

Children’s Hospital Colorado, University of Colorado Anschutz Medical Campus: Katja M. Gist, Imran A Sayed, John Brinton, Larisa Strom

Children’s Hospital of Philadelphia: Kathleen Chiotos, Allison M. Blatz, Giyoung Lee, Ryan H. Burnett, Guy I. Sydney, Danielle M. Traynor

Cox Medical Center Springfield: Steven K. Daugherty, Sam Atkinson, Kelly Shrimpton

Detar Family Medicine residency: Sidney Ontai, Brian Contreras, MD, Uzoma Obinwanko, Nneka Amamasi, Amir Sharafi

Detroit Medical Centre: Sarah Lee, Zahia Esber, Chetna Jinjvadia

George Washington University: David P. Yamane, Ivy Benjenk, Nivedita Prasanna

Howard University Hospital: Orma Smalls

Jacobs Medical Center UC San Diego Health – La Jolla: Atul Malhotra, Abdurrahman Husain, Qais Zawaydeh

JPS Health Network: Steven Q. Davis, Valentina Jovic, Valentina Jovic, Max Masuda, Amanda Hayes

Lakes Region General Hospital: Michael Smith, William Snow, Riley Liptak, Hannah Durant, Valerie Pendleton, Alay Nanavati, Risa Mrozowsk

LifeBridge Health/Sinai and Northwest Hospitals: Namrata Nag, Jeff Brauer, Ashwin Dharmadhikari, Sahib Singh, Franco Laghi, Ghania Naeem, Andrew Wang, Kevin Bliden, Amit Rout, Jaime Barnes, Martin Gesheff, Asha Thomas, Melbin Thomas, Alicia R. Liendo, Jovan Milosavljevic, Kenan Abbasi, Nicholas B. Burley, Nicole Rapista, Samuel Amankwah, Sanjay K Poudel, Saroj Timilsina, Sauradeep Sarkar, Oluwasayo Akinyosoye, Shashi K. Yalamanchili, Sheena Moorthy, Sonia Sugumar, Jonathan Ford, Martin C. Taylor, Charlotte Dunderdale, Alyssa Henshaw, Mary K. Brunk, Jessica Hagy, Shehryar Masood, Sushrutha Sridhar

Loyola University Medical Center: Yuk Ming Liu, Sarah Zavala, Sarah Zavala, Esther Shim

M Health-Fairview, University of Minnesota: Ronald A. Reilkoff, Julia A. Heneghan, Sarah Eichen, Lexie Goertzen, Scott Rajala, Ghislaine Feussom, Ben Tang

MacNeal Hospital Loyola Medicine: Christine C. Junia, Robert Lichtenberg, Hasrat Sidhu, Diana Espinoza, Shelden Rodrigues, Maria Jose Zabala, Daniela Goyes, Ammu Susheela, Buddhi Hatharaliyadda, Naveen Rameshkumar, Amulya Kasireddy, Genessis Maldonado, Lisseth Beltran, Akshata Chaugule, Hassan Khan

Mayo Clinic Arizona: Rodrigo Cartin-Ceba, Ayan Sen, Amanda Palacios, Giyth M. Mahdi

Mayo Clinic Rochester: Rahul Kashyap, Ognjen Gajic, Vikas Bansal, Aysun Tekin, Amos Lal, John C. O’Horo, Neha N. Deo, Mayank Sharma, Shahraz Qamar, Cory J. Kudrna, Juan Pablo Domecq

Mayo Clinic, Eau Claire: Abigail T. La Nou, Marija Bogojevic

Mayo Clinic, Florida: Devang Sanghavi, Pramod Guru, Karthik Gnanapandithan, Hollie Saunders, Zachary Fleissner, Juan Garcia, Alejandra Yu Lee Mateus, Siva Naga Yarrarapu

Mayo Clinic, Mankato: Syed Anjum Khan, Nitesh Kumar Jain, Thoyaja Koritala

Medical Center Health System, Odessa: Alexander Bastidas, Gabriela Orellana, Adriana Briceno Bierwirth, Eliana Milazzo, Juan Guillermo Sierra, Thao Dang

Medical Center Navicent Health: Amy B. Christie, Dennis W. Ashley, Rajani Adiga

Mercy Hospital and Medical Center, Chicago: Travis Yamanaka, Nicholas A. Barreras, Michael Markos, Anita Fareeduddin, Rohan Mehta

Mercy Hospital, Saint Louis: Chakradhar Venkata, Miriam Engemann, Annamarie Mantese

Millard Fillmore Suburban Hospital: Anna Eschler, Mary Hejna, Emily Lewandowski, Kristen Kusmierski, Clare Martin

Montefiore Medical Center The Bronx: Jen-Ting Chen, Aluko Hope, Zoe Tsagaris, Elise Ruen, Aram Hambardzumyan

OSF Saint Francis Medical Center: Bhagat S. Aulakh, Sandeep Tripathi, Jennifer A. Bandy, Lisa M. Kreps, Dawn R. Bollinger, Jennifer A. Bandy

OSF Saint Francis Medical Center: Bhagat S. Aulakh, Sandeep Tripathi, Jennifer A. Bandy, Lisa M. Kreps, Dawn R. Bollinger, Jennifer A. Bandy

Parkview Health System, Fort Wayne: Roger Scott Stienecker, Andre G. Melendez, Tressa A. Brunner, Sue M Budzon, Jessica L. Heffernan, Janelle M. Souder, Tracy L. Miller, Andrea G. Maisonneuve

Samaritan Health Services: Brian L. Delmonaco, Anthony Franklin, Mitchell Heath

Sarasota Memorial Hospital: Antonia L. Vilella, Sara B. Kutner, Kacie Clark, Danielle Moore

St. Joseph’s Candler Health System: Howard A. Zaren, Stephanie J. Smith, Grant C. Lewis, Lauren Seames, Cheryl Farlow, Judy Miller, Gloria Broadstreet

St.Mary Medical Center, Langhorne: Umang Patel, Jordesha Hodge, KrunalKumar Patel, Shivani Dalal, Himanshu Kavani, Sam Joseph

Stamford Health: Michael A. Bernstein, Ian K. Goff, Matthew Naftilan, Amal Mathew, Deborah Williams, Sue Murdock, RN, Maryanne Ducey, Kerianne Nelson

Stanford Hospital and Clinics: Paul K Mohabir, Connor G O’Brien, Komal Dasani

The Children’s Hospital at OU Medicine: Neha Gupta, Tracy L Jones, Shonda C Ayers, Amy B Harrell, Dr. Brent R Brown

The University of Tennessee Medical Center: Megan Edwards, Caleb Darby, Kristy Page, Amanda Brown, Jessie McAbee

Thomas Jefferson University Hospital: Katherine A. Belden, Michael Baram, Devin M. Weber, Rosalie DePaola, Yuwei Xia, Hudson Carter, Aaron Tolley, Mary Barletta

Truman Medical Centers: Mark Steele, Laurie Kemble

Tulane University Medical Center and University Medical Center New Orleans: Joshua L. Denson, A. Scott Gillet, Margo Brown, Rachael Stevens, Andrew Wetherbie, Kevin Tea, Mathew Moore

UNC Medical Center: Benjamin J Sines, Thomas J Bice

University Medical Center of Southern Nevada Las Vegas; University of Nevada, Las Vegas: Rajany V. Dy, Alfredo Iardino, Jill Sharma, Julia Christopher, Marwan Mashina, Kushal Patel

University of Alabama at Birmingham: Erica C. Bjornstad, Nancy M. Tofil, Scott House, Isabella Aldana

University of Arkansas for Medical Sciences: Nikhil K. Meena, Jose D. Caceres, Nikhil K Meena, Sarenthia M. Epps, Harmeen Goraya, Kelsey R. Besett, MD, Ryan James, Lana Y. Abusalem, Akash K. Patel, Lana S Hasan

University of Cincinnati: Dina Gomaa B.S., Michael Goodman, Devin Wakefield, Anthony Spuzzillo, John O. Shinn II

University of Iowa Carver College of Medicine: Patrick W. McGonagill, Colette Galet, Janice Hubbard, David Wang, Lauren Allan, Aditya Badheka, Madhuradhar Chegondi

University of Kansas Medical Center: Usman Nazir, Garrett Rampon, Jake Riggle, Nathan Dismang

University of Louisville Hospital: Ozan Akca, Rainer Lenhardt, Rodrigo S. Cavallazzi, Ann Jerde, Alexa Black, Allison Polidori, Haily Griffey, Justin Winkler, Thomas Brenzel

University of Michigan Health System: Pauline Park, Andrew Admon, Sinan Hanna, Rishi Chanderraj, Maria Pliakas, Ann Wolski, Jennifer Cirino

University of Missouri, Columbia: Dima Dandachi, Hariharan Regunath, Maraya N. Camazine, Grant. E. Geiger, Abdoulie O. Njai, Baraa M. Saad

University of Vermont Larner College of Medicine: Renee D. Stapleton, Anne E. Dixon, Olivia Johnson, Sara S. Ardren, Stephanie Burns, Anna Raymond, Erika Gonyaw, Kevin Hodgdon, Chloe Housenger, Benjamin Lin, Karen McQuesten, Heidi Pecott-Grimm, Julie Sweet, Sebastian Ventrone

Wake Forest University School of Medicine; Wake Forest Baptist Health Network: Ashish K. Khanna, Lynne Harris, Bruce Cusson, Jacob Fowler, David Vaneenenaam, Glen McKinney, Imoh Udoh, Kathleen Johnson

Yale New Haven Health New Haven: Kevin Sheth, Abdalla Ammar, Mahmoud Ammar, Victor Torres Lopez, Charles Dela Cruz, Akhil Khosla, Samir Gautam

## Notes

Financial support and sponsorship: **None**

The VIRUS Registry is supported, in part, by the Gordon and Betty Moore Foundation, and Janssen Research & Development, LLC

### Competing Interest Statement

The authors have declared no competing interest.

### Clinical Trial

NCT04486521

### Funding Statement

No external funding was provided for this study.

### Author Declarations

The study is approved by the Ethics Committee (office of research affair at King Faisal Specialist Hospital Project ID 2201053), The Society of Critical Care Medicine (SCCM) Scientific Review Committee, and SCCM Discovery Steering Committee. In some participating hospitals, verbal consent was obtained whereas in other countries the requirement for patient consent was waived by local research ethics committees. Local investigators were responsible for obtaining local approval in line with applicable regulations.

### Summary of Updates

We added revised section for the statistical analysis, results and discussion section based on reviewers comments

